# A Unified Multiplex Digital PCR-based Platform for Multi-class Biomarker Quantification from Low-volume Blood

**DOI:** 10.1101/2025.08.31.25334810

**Authors:** Jiumei Hu, Joon Soo Park, Hanran Lei, Patarajarin Akarapipad, Jialin Sun, Dong Jin Park, Weiwen Cui, Tza-Huei Wang

## Abstract

Simultaneous quantification of multiple biomarker classes from blood is essential for improving diagnostic accuracy, yet current methods remain limited by low multiplexing capacity, reliance on separate instruments and workflows, and inconsistent signal readouts that hinder data integration. Here, we present FluoMag-dCoDe (<u>Fluo</u>rescence-coded, <u>Mag</u>netic bead-enhanced <u>d</u>igital <u>Co</u>-<u>De</u>tection of multi-analytes), a unified platform that converts diverse analytes into distinct fluorescence-coded DNA templates for multiplexed quantification in a single digital PCR (dPCR) reaction using a universal primer and TaqMan probe set. We demonstrate co-measurement of eight mRNA and three protein immune-response biomarkers in a two-color dPCR format. The platform achieves high quantitative fidelity with minimal cross-interference, showing strong concordance with single-plex RT-qPCR for mRNAs and superior analytical sensitivity to ELISA for proteins – including reliable detection of TRAIL where ELISA failed – from ∼ 35 µL fingerstick-compatible blood volumes. By unifying mRNA and protein quantification in a single standardized readout while sustaining robust performance from low blood volumes, FluoMag-dCoDe establishes a scalable approach for high-dimensional biomarker analysis in precision diagnostics.

## Introduction

Biomarkers, defined as measurable substances or characteristics that serve as indicators of physiological or pathological processes, are fundamental tools in modern diagnostics and personalized medicine.^1^ Among the diverse range of potential biomarkers, nucleic acids and proteins have emerged as particularly powerful due to their inherent specificity, sensitivity, and amenability to high-throughput analysis.^2–7^ While single biomarkers are often employed for disease diagnosis, utilizing a panel of biomarkers has demonstrated a significant improvement in diagnostic accuracy by better reflecting the complexity and heterogeneity of biological systems.^8,9^ Moreover, given that biological systems are regulated through highly interconnected networks involving diverse biomolecules, the simultaneous analysis of multiple classes of biomarkers would offer a more comprehensive and accurate representation of disease states and physiological conditions. Indeed, several studies have demonstrated enhanced diagnostic sensitivity when multiple classes of biomarkers are assessed together.^10, 11^

Existing *in vitro* diagnostic tests, such as polymerase chain reaction (PCR) and immunoassays, are primarily developed to detect only a single class of biomarker. As a result, comprehensive analysis of multiple analyte classes (multi-analytes) typically requires separate workflows and instrumentation, significantly increasing the complexity and cost of diagnostic procedures. The lack of integration across different assay types can introduce variability and impede the standardization of multi-analyte diagnostic approaches.^12, 13^ To overcome such limitations, a critical need exists for unified platforms capable of co-quantifying multi-analytes using a single signal generation and detection system. Such platforms must provide accurate and sensitive analyses while minimizing workflow complexity and instrumentation requirements.^14–16^

Significant efforts have been dedicated to developing multi-analyte detection platforms capable of simultaneously detecting nucleic acids and proteins for disease diagnostics, such as cancer^17, 18^ and infectious diseases.^19–22^ While these methods demonstrate the ability to detect multiple classes of biomolecules, their multiplexing capacity is often restricted to only two to four analytes, limiting their utility for analyzing multiple panels of biomarkers. Moreover, protein detection in these platforms predominantly relies on immunobinding-based signals, resulting in lower detection sensitivity compared to target amplification-based nucleic acid detection.^18–22^ Alternative high-throughput approaches employing protein-to-nucleic acid transformation followed by sequencing-based readouts offer highly multiplexed co-detection of nucleic acids and proteins.^23–26^ However, these approaches often involve complex and time-consuming sample processing steps and require specialized equipment, confining their application to research setting rather than routine diagnostics. Despite significant improvements in multi-analyte detection, there remains an unmet need for a streamlined, multiplexed platform capable of quantifying diverse biomolecule classes with the sensitivity, throughput, and simplicity suitable for diagnostic applications.

In the present work, we introduce <u>Fluo</u>rescence-coded, <u>Mag</u>netic bead-enhanced <u>d</u>igital <u>Co</u>-<u>De</u>tection of multi-analytes (FluoMag-dCoDe), a streamlined multi-analyte detection platform designed for the simultaneous quantification of multiple classes of biomolecules. FluoMag-dCoDe leverages target-specific oligonucleotide extension to convert multiple analytes into distinct fluorescence-coded DNA templates, which are then co-quantified in a single digital PCR (dPCR) reaction using universal primers and TaqMan probes. By generating a unified signal output from multi-analytes, FluoMag-dCoDe overcomes the inherent limitations of conventional methods that rely on separate signal generation setups for each analyte. Multiplexing is facilitated by the fluorescence-coding strategy, where each target is identified and quantified based on a unique combination of multi-color fluorescence intensities determined by the distinct number of TaqMan probe binding sites within each template.^27, 28^ To validate its performance, we demonstrated FluoMag-dCoDe for the quantitative detection of two panels of infection-associated immune-response biomarkers – 8 mRNAs^29^ and 3 proteins^30, 31^ – utilizing only two fluorescence colors. The multiplexed quantification accuracy of FluoMag-dCoDe was validated against the gold-standard singleplex RT-qPCR and ELISA for mRNA and protein, respectively. Furthermore, FluoMag-dCoDe successfully detected multi-analytes from a blood sample as small as 35 µL, a volume compatible with fingerstick collection, demonstrating its potential for practical use in routine diagnostic applications.

## Results

### Assay overview

FluoMag-dCoDe begins with blood sample processing, involving separate RNA extraction and plasma separation steps to enable downstream workflows for detecting multiple mRNA and protein biomarkers. Both workflows convert their respective targets into universal fluorescence-coded DNA templates via target-specific oligonucleotide extension, allowing simultaneous quantification of both analytes in a single fluorescence-coding dPCR reaction using only one set of primers and TaqMan probes (**Figure 1**). Such a single final reaction generates a unified signal output, streamlining multi-analyte data analysis and reducing technical variability. Moreover, the fluorescence-coding strategy in FluoMag-dCoDe facilitates multiplexed target quantification using a limited number of fluorescence channels.

**Figure 1.**
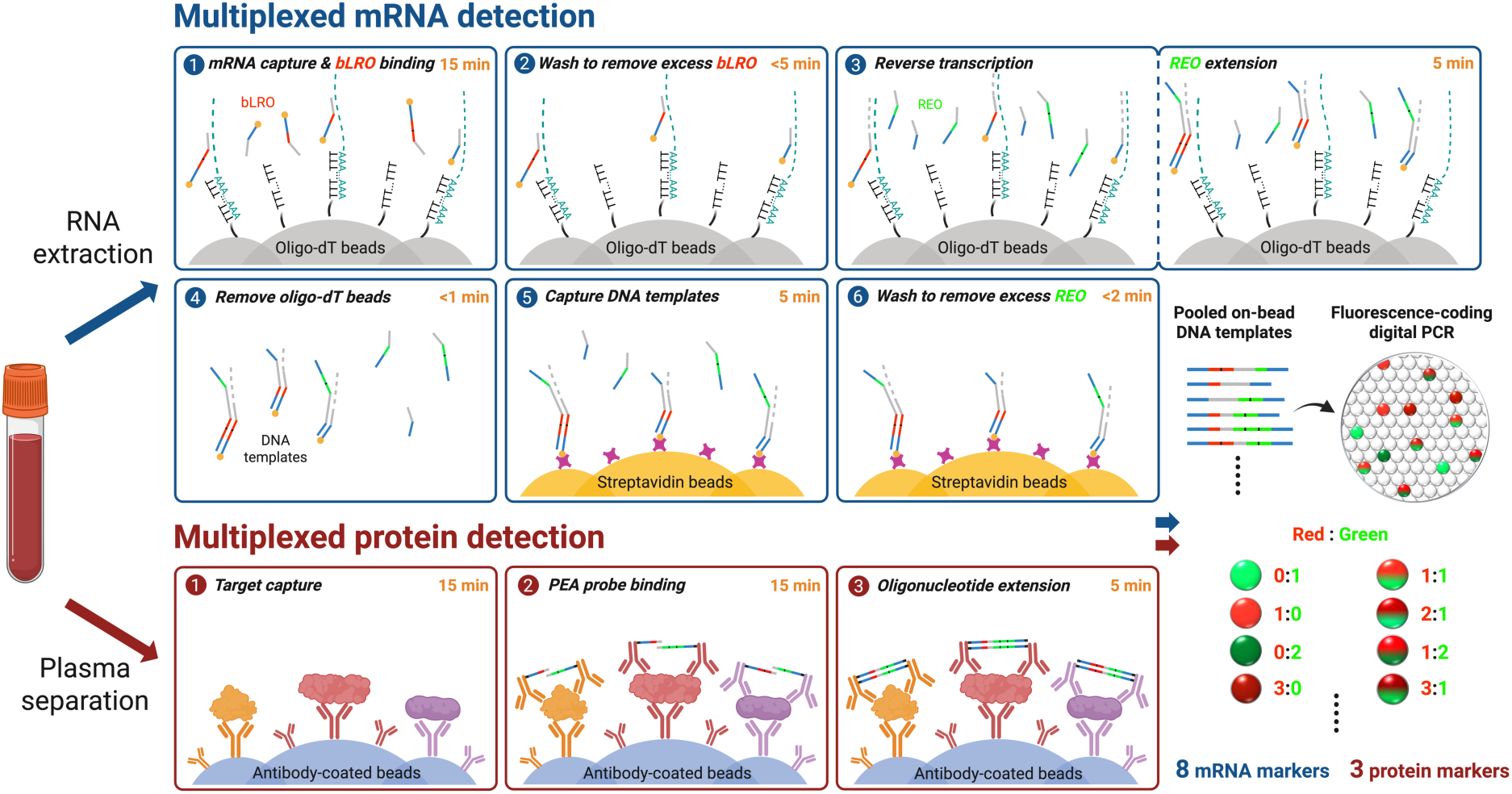
Schematic overview of FluoMag-dCoDe for simultaneous detection of mRNA and protein biomarkers. Blood samples are processed by RNA extraction and plasma separation to enable parallel mRNA and protein detections. For mRNA detection, targets are captured by oligo-dT beads and hybridized with bLROs, followed by one-step RT and REO extension, and digital capture of the fluorescence-coded DNA templates using excess streptavidin beads. For protein detection, proteins are digitally captured by excess antibody-coated beads, recognized by PEA probes, and converted into on-bead fluorescence-coded DNA templates via oligonucleotide extension. Beads from both workflows are then mixed and encapsulated into droplets for one-pot fluorescence-coding ddPCR, where end-point fluorescence intensities are measured to decode and quantify mRNA and protein targets.

For mRNA detection, consecutive reverse transcription (RT) and oligonucleotide extension are employed to generate distinct fluorescence-coded DNA templates for multiple mRNA targets. To achieve this, target-specific oligonucleotide pairs, each consisting of a biotinylated left RT oligonucleotide (bLRO) and a right extension oligonucleotide (REO), are designed with a universal primer binding site and predefined number of TaqMan probe binding sites at their 5’ ends. To prevent excess bLROs and REOs from competing with universal primers during PCR amplification – a phenomenon observed in previous fluorescence-coding strategy that compromised fluorescence signal fidelity^28^ – a two-bead washing system is developed. Initially, mRNA is captured using oligo-dT beads while bLROs selectively bind to their target sites on mRNA. After washing to remove unbound bLROs, the beads are resuspended in an RT-PCR mixture containing REOs, facilitating cDNA synthesis along the bLROs and subsequent REO extension on the cDNA to generate fluorescence-coded DNA templates. These templates, now released from the oligo-dT beads, are then digitally captured using excess streptavidin beads (i.e., one or no template per bead), followed by a final wash to remove free REOs.

For protein measurement, magnetic bead-enhanced proximity extension assay (PEA)^32^ is employed to generate fluorescence-coded DNA templates from multiple protein targets via two sequential immunobinding steps. First, plasma proteins are stochastically captured on an excess of antibody-coated beads, such that most beads carry either one or no protein target. Next, multiple pairs of antibody–oligonucleotide conjugates (thereafter referred to as PEA probes), each designed with unique oligonucleotide sequences for fluorescence-coding, are added to recognize and bind the bead-captured proteins in close proximity. Upon oligonucleotide extension, distinct fluorescence-coded DNA templates are generated on bead surface, each representing a specific protein target.

Both mRNA and protein assay workflows requires less than 40 min to generate the on-bead fluorescence-coded DNA templates. At this step, each template carries a target-specific number and arrangement of probe-binding sites, intrinsically encoding analyte identity that will later appear as distinct fluorescence signatures during droplet dPCR (ddPCR). Next, these beads are partitioned into a large number of discrete droplets for dPCR reactions. During ddPCR amplification, DNA polymerase cleaves the bound TaqMan probes via its 5’-3’ exonuclease activity to generate fluorescence signals. The unique combination of two fluorescent colors, each with different fluorescence intensities, produces a unique fluorescence code for each DNA template. Subsequently, the end-point fluorescence intensities of individual droplets are measured at two fluorescence channels, forming multiple droplet populations characterized by unique fluorescence signatures. By counting the number of droplets in each population, all the mRNA and protein targets are reliably identified and quantified.

### Development of multiplexed mRNA detection system in FluoMag-dCoDe

As a proof of validation of the multiplexed mRNA assay, we selected an eight-gene signature previously identified as a host-response mRNA biomarker panel for differentiating bacterial and viral infections.^29^ This panel includes three genes upregulated in bacterial infections (SMARCD3, ICAM1, EBI3; referred to as “bacterial genes”) and five genes overexpressed in viral infections (HESX1, FCER1A, SUCLG2, IFI27, JUP; referred to as “viral genes”).

We first designed bLRO and REO sequences specific to each target (**Figure 2A** and **Table S1**). Each bLRO contains a biotin tag, a universal forward primer binding site, varying number of HEX TaqMan probe binding sites, and a target-specific binding region. Similarly, each REO comprises a universal reverse primer binding site, varying number of FAM TaqMan probe binding site, and a target-specific cDNA binding region. Using 50 ng of human total RNA, the two-dimensional fluorescence amplitude plots showed that each singleplex assay generated distinct droplet clusters with unique fluorescence signatures in the HEX (red) and FAM (green) channels (**Figure S1**). The observed signatures closely matched those generated from synthetic fluorescence-coded DNA templates corresponding to each mRNA target (**Table S2**), confirming efficient mRNA-to-DNA conversion and reliable fluorescence-coding through PCR amplification (**Figure S2**). When testing varying inputs of human total RNA, the singleplex assays showed a proportional decrease in ddPCR signal output with decreasing RNA input, while maintaining clean background signals (**Figure S3**). Strong linear relationships between RNA input and ddPCR signal output were observed for all eight genes, with an average R^2^ value above 0.95, demonstrating robust mRNA quantification capability of the singleplex format of FluoMag-dCoDe (**Figure 2B**). Furthermore, comparison of Cycle threshold (Ct) values from gold-standard RT-qPCR with the ddPCR signal outputs yielded a Pearson correlation coefficient of 0.75, suggesting good quantitative agreement between the two methods (**Figure 2C**).

**Figure 2.**
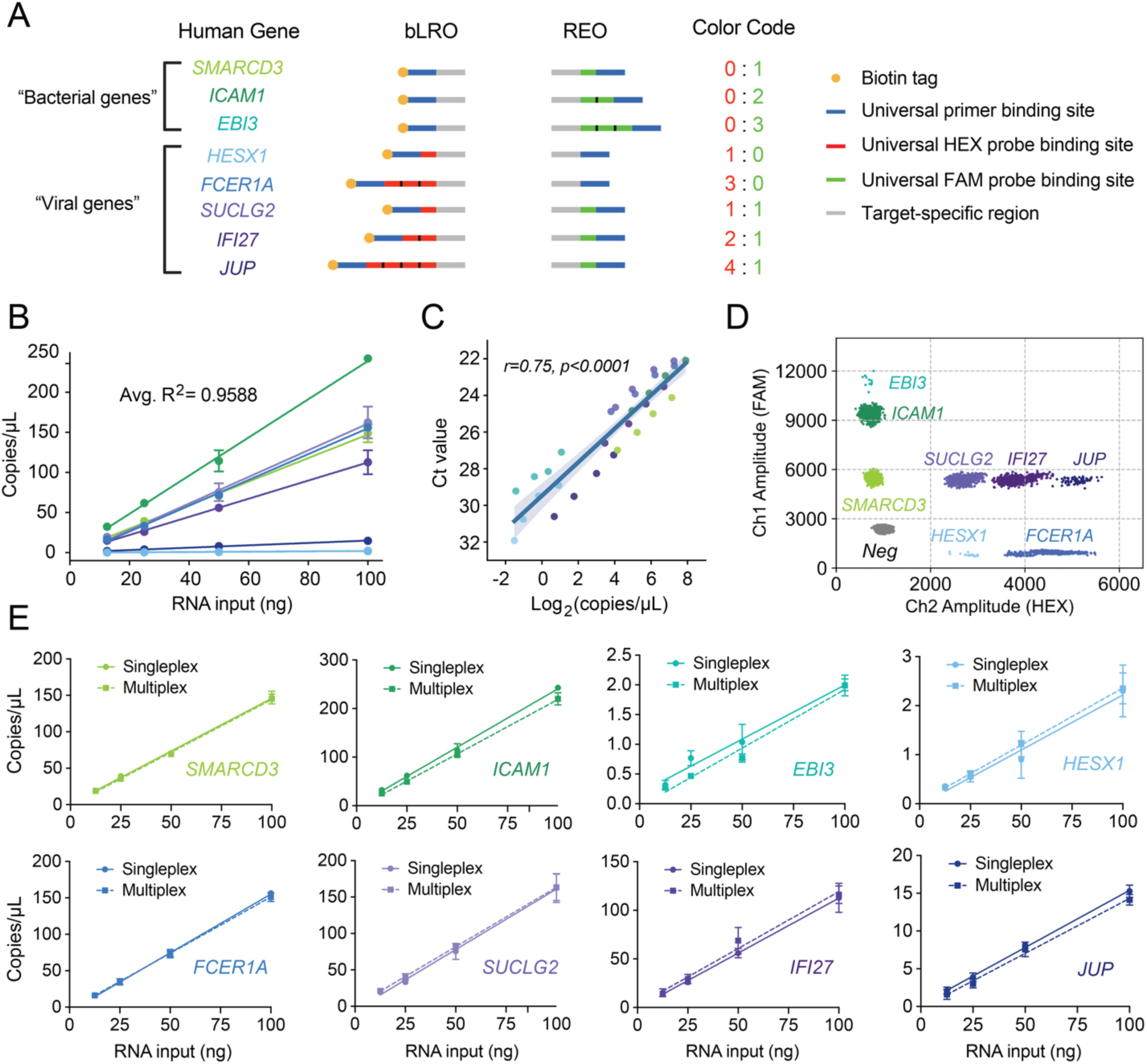
Development of multiplexed mRNA detection system in FluoMag-dCoDe. (A) Fluorescence-codes for each mRNA target, including three “bacterial genes” and five “viral genes”, are designed by varying the number of universal HEX and FAM TaqMan probe binding sites. (B) Aggregated standard curves show good linear relationship between human total RNA inputs and ddPCR signal outputs. Each datapoint represents a mean ± standard deviation from three replicates. (C) Pearson correlation analysis with r=0.75 and p<0.0001 indicates a good correlation between the readouts from FluoMag-dCoDe and those from gold-standard RT-qPCR for each target. The shaded area represents the standard error of the slope estimate. (D) Two-dimensional FAM-versus-HEX amplitude plot of the multiplex reaction (50 ng total RNA) shows eight target-specific droplet populations and a single negative population. (E) Linear regression curves (n=3) from singleplex (solid) and multiplex (dashed) assays for each target show no significant difference in both slopes and intercepts.

We next performed eight-plex detection in a single reaction by incorporating the bLROs and REOs specific to all the mRNA targets. Using 50 ng human total RNA as the input, FluoMag-dCoDe showed eight well-separated droplet populations at distinct locations on the two-dimensional amplitude plot, which corresponded to the pre-defined fluorescence-codes for each target (**Figure 2D**). Comparison of the multiplexed result with the singleplex assays showed high concordance (Student’s t-test p=0.82) in quantifying all eight mRNA targets (**Figure S4**). To further assess the performance of the FluoMag-dCoDe, we tested varying amounts of human total RNA input (100, 50, 25, and 12.5 ng) and consistently observed eight target-specific, fluorescence-coded droplet populations, with the number of positive droplets decreasing proportionally with lower RNA input (**Figure S5**). Comparison of the linear regression curves between the multiplex and singleplex assays using an ANCOVA extra-sum-of-squares F-test showed no significant difference in either slope or intercept across all eight targets, confirming that the multiplex assay maintained quantification accuracy without compromising the detection efficiency of individual targets (**Figure 2E** and **Table S3**). Collectively, these results demonstrate that our assay provides accurate and reliable quantification of multiple mRNA targets in a single reaction, highlighting its effectiveness for multiplexed gene expression analysis.

### Development of multiplexed protein detection system in FluoMag-dCoDe

We selected an established three-protein biomarker panel, including CRP, IP10, and TRAIL, for distinguishing bacterial from viral infections.^33, 34^ CRP is typically elevated in bacterial infections, while IP10 is more specific to viral infections. TRAIL level exhibits a context-dependent pattern, increasing in viral infections and decreasing in bacterial infections.

To generate fluorescence-coding signatures for protein biomarkers distinct from those for mRNA biomarkers, we prepared target-specific PEA probe pairs by conjugating antibodies to oligonucleotides containing unique numbers of universal HEX and FAM probe binding sites (**Figure 3A** and **Table S1**). Using recombinant proteins spiked into chicken plasma as contrived samples, each singleplex assay produced distinct ddPCR fluorescence intensities at both channels that matched the predesigned fluorescence-codes (**Figure S6**). Moreover, by testing a dilution series for each target, the singleplex assay demonstrated reliable quantification across a wide dynamic range, achieving detection limits of 360 fg/mL for CRP, 800 fg/mL for IP10, and 7.5 fg/mL for TRAIL (**Figure S7**), sensitivities that are sufficient in clinically relevant settings and represent ∼5 to 1000-fold improvement over traditional ELISA.

**Figure 3.**
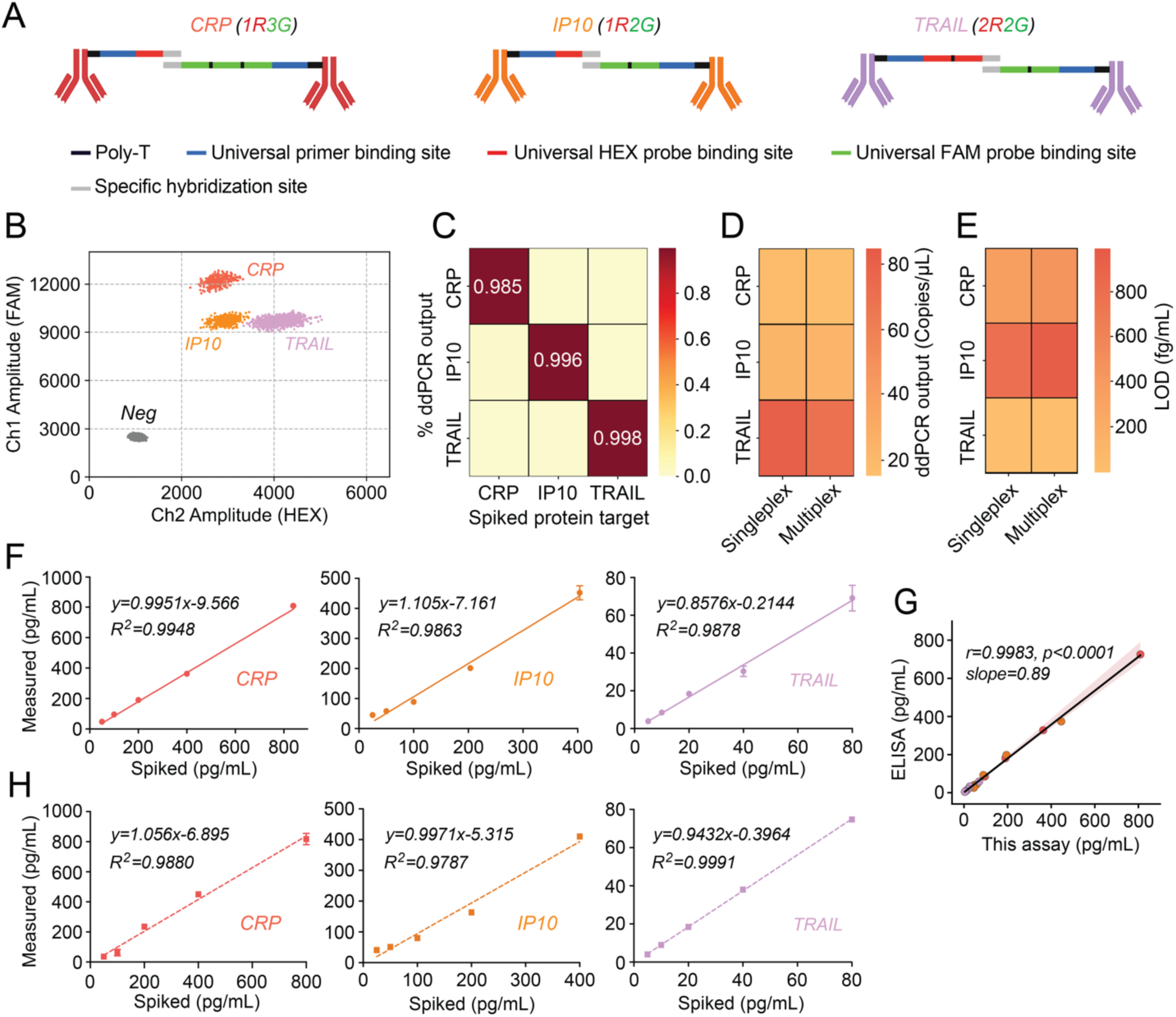
Development of multiplexed protein detection system in FluoMag-dCoDe. (A) Fluorescence-codes for each protein target are generated by conjugating the antibodies to oligonucleotides that carry distinct number of HEX and FAM TaqMan probe binding sites. (B) FAM-versus-HEX amplitude plot from the triplex assay (100 pg/mL CRP and IP10; 1 pg/mL TRAIL) shows three target-specific droplet populations and a negative population. (C) Specificity was tested by spiking one target per condition (same target concentrations as in panel B) while including all bead types and PEA probes. Each condition produced predominate signals only for the spiked target, indicating minimal cross-reactivity. (D) Singleplex and multiplex assays (same target concentrations as in panel B) generated comparable signal output for each target. (E) The singleplex and multiplexed assays demonstrated a similar detection limit for each target. (F) Spiked-versus-measured plots for each singleplex assay showed high quantification accuracy of these biomarkers in human plasma using the UniQ Buffer as a sample diluent. (G) Strong concordance on the quantification results obtained from each singleplex assay and those from ELISA was observed via Pearson correlation analysis. The shaded area represents the standard error of the slope estimate. (H) Spiked-versus-measured plots for each targets in the multiplexed assay format further underscore the suitability of using UniQ Buffer for quantitative measurement of the three biomarkers simultaneously in human plasma.

Next, we evaluated the capability of FluoMag-dCoDe for simultaneously measuring all three protein targets in a single reaction. A mixture of the proteins was spiked into chicken plasma, and the full set of antibody-coated beads along with their corresponding PEA probes was introduced during the immunobinding steps. The result showed distinct droplet populations that represent each target, confirming the reliability of the designed fluorescence-codes for multiplexed protein biomarker detection (**Figure 3B**). We further performed a systematic evaluation of the triplex protein assay, focusing on specificity, quantification accuracy, and detection sensitivity. Specificity was examined by individually spiking only one protein target into chicken plasma while using the beads and PEA probes for all three targets per reaction. The resulting ddPCR readouts showed predominant signal only for each spiked protein, indicating minimal cross-reactivity among multiple targets (**Figure 3C**). To evaluate quantification accuracy, we compared ddPCR readouts obtained in the triplex assay with those from target-matched singleplex reactions. As shown in **Figure 3D**, multiplex signals were statistically indistinguishable (Student’s t-test p=0.89) from their singleplex counterparts, suggesting uncompromised signal generation in the multiplexed setup. Meanwhile, by testing a dilution series of protein mixtures, the triplex assay achieved sensitivities comparable to those observed in singleplex assays, with detection limits down to 450, 970, and 11 fg/mL for CRP, IP10, and TRAIL, respectively (**Figure 3E** and **Figure S8**). Overall, these results establish FluoMag-dCoDe as a robust platform for sensitive, specific, and multiplexed protein quantification with high analytical precision.

### Multiplexed protein detection in human plasma

To enable precise quantification of protein biomarkers under clinically relevant conditions, we next sought to optimize the triplex assay for use with human plasma. Due to its high content of enzymes, lipids, and other interfering substances, plasma presents a challenging matrix that can disrupt antibody-antigen binding and compromise assay precision, a phenomenon referred to as the matrix effect.^35, 36^ As a result, traditional immunoassays often require dilution of plasma samples with appropriate diluent to mitigate the matrix effect, albeit at the cost of reduced sensitivity.^37^ Leveraging the high analytical sensitivity of FluoMag-dCoDe, we implemented a ≥10-fold dilution to reduce matrix-associated artifacts while preserving reliable quantification of all target proteins. To identify a sample diluent that enables accurate measurement of all three protein biomarkers in diluted human plasma, we systematically compared three commercially available formulations that have been used in bead-based immunoassays, including the StartingBlock Blocking Buffer (SBB),^38^ the Universal Assay Buffer (UAB, utilized in the Luminex system),^39, 40^ and the Quanterix Sample Diluent (QSD, utilized in the SiMoA platform).^38, 41, 42^ We conducted a spike-and-recovery test by spiking equal amounts of protein targets into diluted human plasma with each diluent, while using non-spiked diluted human plasma as a baseline control to account for endogenous protein levels. A recovery rate was then calculated as the ratio of baseline-subtracted signal output obtained from spiked plasma to those from sample diluent, with values between 80% to 120% indicating good detection accuracy.^43^ Our results showed that SBB yielded poor recovery for all three targets, whereas both UAB and QSD achieved good recovery for CRP and TRAIL but showed suboptimal performance for IP10 (**Figure S9**). Notably, the IP10 recovery rates exhibited opposing trends between UAB and QSD, prompting us to investigate whether a blended formulation could offset their respective limitations. Among various mixing ratios, a combination of 70% UAB and 30% QSD, designated as the UniQ Buffer, demonstrated good detection recovery for IP10 (**Figure S10**). To further validate the UniQ Buffer as a sample diluent, we performed spike-and-recovery tests for all three protein targets at high, medium, and low spike-in concentrations within clinically relevant ranges. The results confirmed good recoveries for all targets across the tested conditions (**Figure S11**).

To further validate the quantification accuracy of targets in plasma using the UniQ Buffer, we performed a spiked-and-measured test that could provide quantitative agreement between known input concentrations and those inferred from assay measurement. A linearly-fitted standard curve was first generated between spiked target concentrations in the UniQ Buffer and ddPCR signal readouts across clinically relevant range. We then spiked varying concentrations of target proteins into diluted human plasma, measured the ddPCR signals, and back-calculated the protein levels from the UniQ Buffer-derived standard curves. For each singleplex assay, the resulting spiked- versus-measured plots exhibited strong linearity, with tightly constrained slopes (0.8 to 1.2) and minimal intercepts, indicating high accuracy of quantification using the UniQ Buffer (**Figure 3F**). Comparison on the quantification results for each target obtained from our assay to those measured using gold-standard ELISA also demonstrated a high degree of concordance between the two methods (**Figure 3G**). Moreover, the high quantification accuracy was preserved in the multiplexed assay format, as reflected by well-fitted spiked-versus-measured curves shown in **Figure 3H**. Overall, these results confirm the suitability of the UniQ Buffer for quantitative analysis of these biomarkers in human plasma samples.

### Co-detection of mRNA and protein biomarkers in a single ddPCR reaction

Having established the mRNA and protein detection systems of FluoMag-dCoDe, we next sought to integrate them into a single ddPCR workflow for multiplexed, multi-analyte biomarker analysis. To assess cross-assay compatibility of mRNA and protein detection system, beads carrying fluorescence-coded DNA templates from both systems were pooled together and partitioned into droplets for ddPCR amplification. As shown in **Figure 4A**, the droplet populations representing each target were clearly identified, demonstrating the effectiveness of FluoMag-dCoDe for simultaneous quantification of the mRNA and protein biomarkers with the fluorescence-coding scheme.

**Figure 4.**
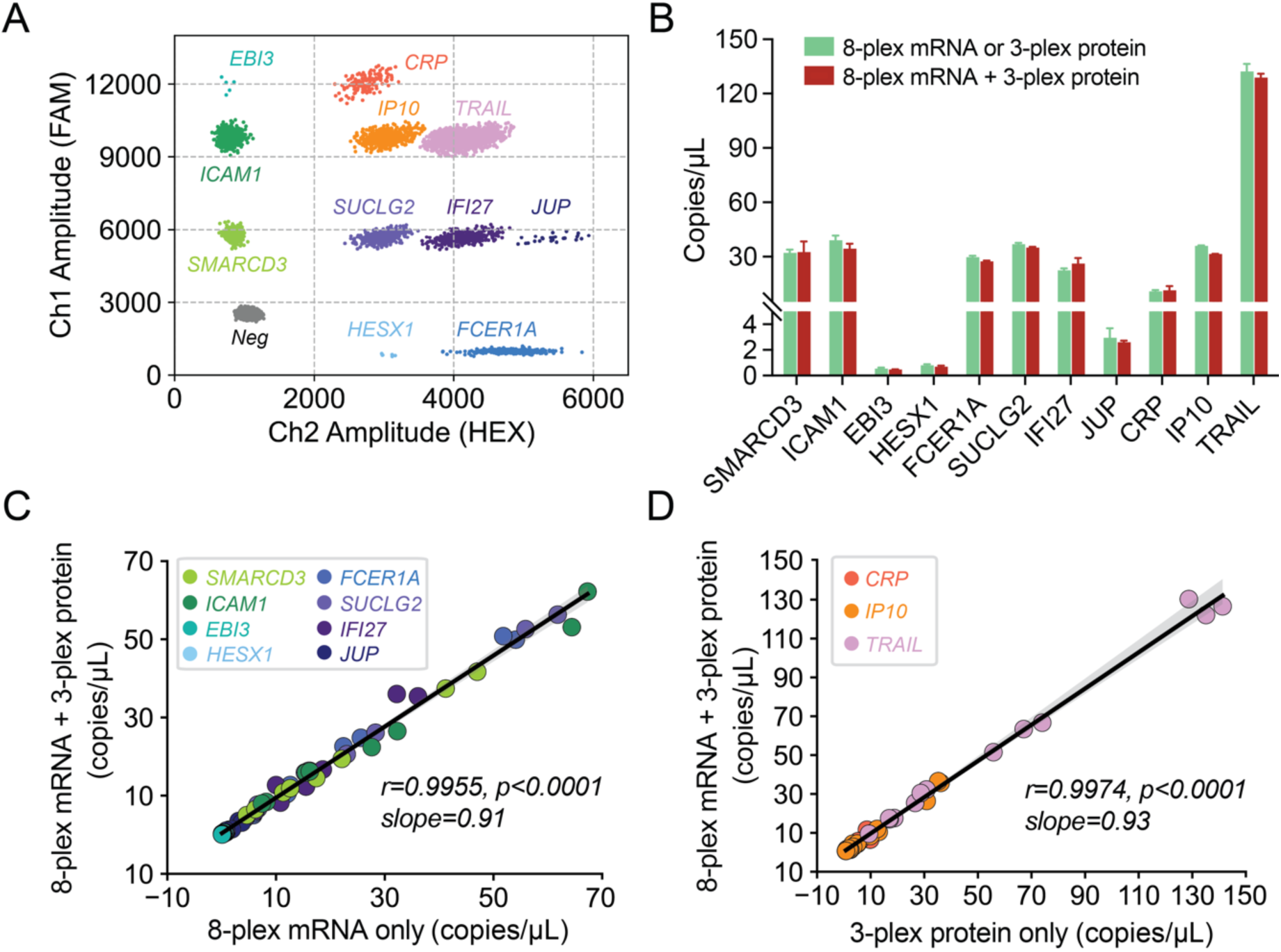
Multiplexed, multi-analyte biomarker detection in a single ddPCR reaction. (A) Two-dimensional amplitude plot resolving eight mRNA and three protein biomarkers in one ddPCR reaction. Inputs: 25 ng human total RNA; CRP 1000 pg/mL, IP10 500 pg/mL, TRAIL 100 pg/mL (spiked in UniQ Buffer). (B) Comparison of ddPCR signal between the integrated assay and individual mRNA or protein assays at identical target inputs as in panel (A), demonstrating preserved quantification accuracy (n=3). (C) Pearson correlation of mRNA signals from the integrated versus mRNA-only assay across varying human total RNA inputs of 100, 50, 25, and 12.5 ng. The integrated assay contains the fixed protein concentration identical to panel (A). (D) Pearson correlation of protein signals for two-fold serial dilutions (CRP: 1000–62.5 pg/mL, IP10: 500–31.25 pg/mL, TRAIL: 100–6.25 pg/mL) between the integrated and protein-only assay. The integrated assay contains a fixed human total RNA input of 25 ng. For panel (C) and (D), each data point represents one reaction. The shaded area represents the standard error of the slope estimate.

To evaluate the quantification accuracy of FluoMag-dCoDe for multi-analytes, we compared ddPCR readouts from the integrated reaction to those obtained from individual single-analyte assays (i.e., 8-plex mRNA assay and 3-plex protein assay) at a fixed target input. The integrated reaction produced signal outputs comparable to those from the single-analyte assays, indicating preserved quantification accuracy across all eleven targets (**Figure 4B**). To further investigate potential cross-interference between mRNA and protein detection in the integrated ddPCR reaction, we varied the input of one analyte type while keeping the other constant. The resulting ddPCR readouts were then compared to those from corresponding single-analyte assays. When varying the input of human total RNA while maintaining fixed protein concentrations, a strong correlation of ddPCR readouts for each mRNA target was observed between the integrated and mRNA-only assay (**Figure 4C**). Similarly, varying concentrations of protein targets with a fixed human total RNA input produced ddPCR readouts comparable to those from the protein-only assay (**Figure 4D**). Collectively, these results confirm that the presence of one analyte type has minimal impact on the detection of the other, demonstrating the high compatibility of mRNA and protein analysis within a single ddPCR reaction and underscoring the robustness of FluoMag-dCoDe as a multiplexed, multi-analyte assay for concurrent mRNA and protein biomarker quantification.

### Multiplexed, multi-analyte biomarker detection from low-volume human blood

In a clinical context, multi-analyte detection platform should be able to measure multiple classes of biomarker from a given specimen. To this end, we evaluated the performance of FluoMag-dCoDe for concurrent detection of host-response transcripts and proteins from whole blood samples. Specifically, blood samples were processed through two parallel streams: one underwent RNA extraction for bead-enhanced RT and extension to produce mRNA-derived universal DNA templates, while the other was subjected to plasma separation for PEA to generate protein-derived universal DNA templates (**Figure 1**). These templates were then combined and analyzed via a one-pot fluorescence-coding ddPCR, yielding discrete, quantitative signals for each biomarker.

Leveraging the high sensitivity of FluoMag-dCoDe (detection limits at the low-nanogram for mRNA and at the fg/mL-level for protein), we aimed to push the limits of low-volume sampling from a single whole blood sample. To preserve high-quality transcripts for mRNA analysis, whole blood was drawn into RNA-stabilizing reagents prior to total RNA extraction. Our optimized workflow for small blood input showed RNA yield proportional to the input volume, with an average yield of ∼40 ng total RNA from only 10 μL of blood (**Figure 5A**). For protein, we evaluated the feasibility of plasma separation from low-volume blood samples using a centrifugation-based method. This approach effectively separated plasma from cellular components, as demonstrated in **Figure S12**. Across all tested volumes, including 200 μL, 100 μL, 50 μL, and 25 μL, the plasma recovery yield remained consistent at approximately 50% of the whole blood volume (**Figure 5B**). Furthermore, spectrophotometric comparison of absorbance at 540 nm (A_540_) between the separated plasma and commercially sourced plasma confirmed high purity, indicating suitability for downstream applications (**Figure S13**). Together, these results demonstrate the feasibility of obtaining high-quality RNA and plasma from small blood quantities, supporting efficient use of limited sample volumes in our dual-modality analysis workflows.

**Figure 5.**
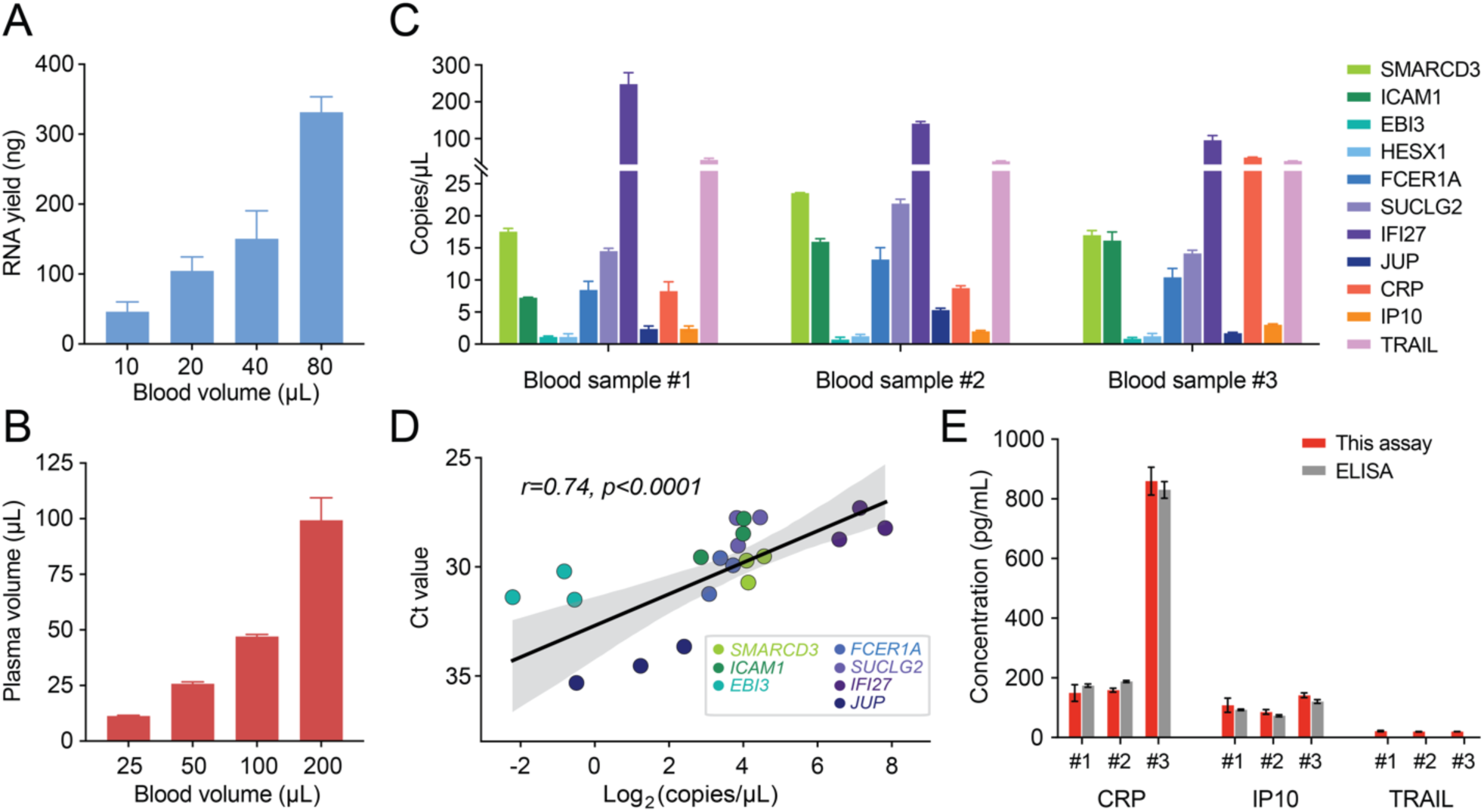
Performance of the eleven-plex FluoMag-dCoDe from low-volume blood samples. (A) Total RNA yield from whole blood samples (n=3) with varying input volumes using an optimized extraction protocol. (B) Plasma recovery volumes as a function of blood input volumes from multiple samples (n=3) following centrifugation-based separation. (C) Fluorescence-coding ddPCR outputs for each biomarker across three blood samples using 10 μL and 25 μL blood for mRNA and protein detections, respectively. Each sample was tested three times. (D) Correlation analysis between mRNA quantification results from FluoMag-dCoDe and those from single-plex RT-qPCR show good concordance. Each datapoint refers to the average value from three replicates for one blood sample. The shaded area represents the standard error of the slope estimate. (E) Protein quantification comparison between FluoMag-dCoDe and ELISA across three blood samples. TRAIL was not detectable in ELISA due to limited sensitivity.

We next validated the performance of the eleven-plex FluoMag-dCoDe, using 10 μL whole blood for mRNA and 25 μL for protein – volumes compatible with fingerstick collection. All mRNA and protein targets were consistently detected across multiple blood samples (**Figure 5C** and **Figure S14**), demonstrating the assay’s robustness for multiplexed, multi-analyte analysis from small blood volumes. To assess quantification accuracy, we further compared our results with gold-standard methods: RT-qPCR for mRNA and ELISA for protein detection. While FluoMag-dCoDe reliably detected all mRNA targets, RT-qPCR failed to detect HESX1 due to limited sensitivity. Among the mRNAs quantified by both methods, a good concordance in quantification between two methods demonstrates the accuracy of FluoMag-dCoDe for quantitative analysis of multiple mRNA targets (r=0.74, **Figure 5D**). Likewise, while FluoMag-dCoDe successfully detected all three protein targets, ELISA detected only CRP and IP10, and failed to detect TRAIL due to limited sensitivity. For the two proteins measurable by both methods, FluoMag-dCoDe showed highly consistent quantification results with those from ELISA (Student’s t-test p=0.98, **Figure 5E**). Overall, these results highlight the robust performance of eleven-plex FluoMag-dCoDe for the simultaneous detection of mRNA and protein biomarkers from small blood volumes, supporting its potential utility in infectious disease diagnostics and broader biomedical research.

## Discussion

FluoMag-dCoDe is a streamlined multi-analyte analysis platform capable of simultaneously quantifying multiple classes of biomolecules with a unified signal output. While the present study focused on mRNAs and proteins, the platform is readily adaptable to a broader range of biomolecules. This versatility is achieved by converting different types of analytes into DNA surrogates^44–47^, enabling the detection of trace levels of target molecules via PCR amplification-based method. Such conversion significantly enhances the analytical sensitivity of non-DNA targets, which traditionally suffer from poor detection limits. Additionally, the unified signal output in FluoMag-dCoDe is generated by a single set of universal primers and probes, simplifying the new assay development and substantially reducing the risk of non-specific interaction often associated with multiple primer sets.

With fluorescence-coding strategy for multiplexing, FluoMag-dCoDe leverages dPCR for individual analysis of fluorescence-coding signature at single-molecule resolution. While the present study demonstrated an eleven-plex multi-analyte analysis using ddPCR with only two fluorescence channels, fluorescence-coding strategy is inherently flexible and not confined to any specific dPCR platform: Both droplet- and array-based^28^ dPCR platforms have been employed, suggesting potential of further improvement with advanced dPCR technologies.^48–51^ A higher degree of multiplexing can be readily accomplished by assigning additional fluorescence-codes to universal PCR templates and integrating additional fluorescence channels. Moreover, the magnetic bead-enhanced approach utilized in FluoMag-dCoDe ensures precise fluorescence-coding by removing unextended oligos – previously shown to interfere with fluorescence signature fidelity^28^ – and thus minimizing background signals for high sensitivity.

FluoMag-dCoDe in the present study was designed to simultaneously quantify two distinct panels of infection-associated immune-response biomarkers – 8 mRNAs^29^ and 3 proteins^30, 31^ – within a single assay. These panels were independently developed to assess host-response biomarkers for distinguishing bacterial and viral infections. While both panels demonstrated diagnostic utility, each panel exhibited complementary diagnostic strengths^52^: the protein panel demonstrated higher sensitivity for identifying bacterial infections, whereas the mRNA panel provided higher specificity. In contrast, for viral infections, the mRNA panel achieved greater sensitivity with specificity comparable to that of the protein panel. Such differential performance indicates the clinical value of integrating mRNA and protein panels for enhanced overall diagnostic performance. Furthermore, the mRNA panel used in this study^29^ represents an upgraded version with improved global applicability, addressing limitations of previous panel^53^ on distinguishing intracellular bacterial and viral infection. Altogether, the integration of these refined host-response biomarker panels using FluoMag-dCoDe underscores its strong potential as a next-generation diagnostic tool for infectious disease differentiation.

In conclusion, FluoMag-dCoDe represents a significant advance in multi-analyte quantification, offering a highly sensitive, multiplexed, and adaptable platform for complex biological analysis from minimal sample volumes. While demonstrated here in the context of infectious disease diagnostics, the versatility and assay development simplicity of FluoMag-dCoDe make it broadly applicable to other fields, such as oncology^10^ and autoimmune diseases.^54^ Future efforts will focus on expanding the range of detectable analytes, increasing multiplexing capacity, and validating clinical performance of FluoMag-dCoDe for diverse diagnostic applications. These ongoing developments hold promise for transforming FluoMag-dCoDe into a powerful tool for precision medicine and global health diagnostics.

## Materials and methods

### Materials

All DNA oligonucleotides used in this study (**Table S1**) including biotinylated LROs, REOs, thiol-terminated PEA probe oligonucleotides, PCR primers, one-step RT-PCR primers, TaqMan probes, and synthetic DNA were purchased from Integrated DNA Technologies, Inc. (Coralville, IA, USA). All the reagents and consumables used for ddPCR including DG8 Cartridges DG8 Gaskets, Droplet Generation Oil for Probes, ddPCR 96-Well Plates, Pieceable Foil Heat Seal, and 2× ddPCR Supermix for Probes (No dUTP) were purchased from Bio-Rad Laboratories, Inc (Philadelphia, PA, USA). Lithium chloride solution (LiCl), Lithium dodecyl sulfate (LiDS), Sodium chloride (NaCl), and Tween 20 were purchased from MilliporeSigma (St. Louis, MO, USA). TaqMan Lyo-ready 1-step RT-qPCR Master Mix, Dithiothreitol (DTT), EDTA, Tempus™ Blood RNA Tube, Tempus^TM^ Spin RNA Isolation Kit, Qubit RNA HS Assay Kit, Human CRP ELISA Kit, Human IP10 ELISA Kit, Human TRAIL ELISA Kit, StartingBlock Blocking Buffer, and Universal Assay Buffer were purchased from ThermoFisher Scientific (Waltham, MA, USA). Recombinant Human CRP/IP10/TRAIL Protein, Human CRP/IP10/TRAIL Polyclonal Antibody were purchased from R&D Systems (Minneapolis, MN, USA). The Human Blood Peripheral Leukocytes Total RNA was purchased from Takara Bio USA, Inc. (Mountain View, CA, USA). Oligo d(T)25 Magnetic Beads and Dynabeads™ M-280 Streptavidin Beads were purchased from New England BioLabs, Inc. (Ipswich, MA, USA) and ThermoFisher Scientific, respectively. The Quanterix Sample Diluent was purchased from Fisher Scientific (Hampton, NH, USA). Chicken plasma in sodium EDTA was purchased from Rockland Immunochemicals, Inc (Pikesville, MD, USA). Human Whole Blood K2 EDTA was purchased from BioIVT (Hicksville, NY, USA). Tris-HCl and DEPC-treated water were purchased from Quality Biological Inc. (Gaithersburg, MD, USA). Evagreen dye was purchased from Biotium (Hayward, CA, USA). A complete list of reagents utilized in the magnetic bead-enhanced PEA procedure for protein detection is available in our previously published paper.^32^

### mRNA detection

For each singleplex reaction, 2 μL of oligo-dT beads were first washed twice with 80 μL of Lysis/Binding buffer (100 mM Tris-HCl, 1 mM EDTA, 0.5 M LiCl, 0.5% LiDS, 5 mM DTT, 0.05% Tween-20). The beads were then resuspended in 50 μL of Lysis/Binding buffer containing bLRO at a final concentration of 500 nM and human total RNA. The mixture was heated at 65°C for 5 min and then cooled to 4°C for 1 min to minimize the impact of mRNA secondary structure on binding. Next, the beads were incubated at room temperature on an end-over-end rotator for 10 min to facilitate mRNA capture and the binding of bLRO onto mRNA targets. The beads were then sequentially washed as follows: twice with 80 μL of Wash Buffer I (20 mM Tris-HCl, 1 mM EDTA, 0.5 M LiCl, 0.1% LiDS, 5 mM DTT, 0.05% Tween-20), twice with 80 μL of Wash Buffer II (20 mM Tris-HCl, 1 mM EDTA, 0.5 M LiCl, 0.05% Tween-20), and once with 80 μL of Low-Salt Buffer (20 mM Tris-HCl, 1 mM EDTA, 0.2 M LiCl, 0.05% Tween-20). The washed beads were resuspended in 10 μL of TaqMan Lyo-Ready 1-Step RT-qPCR buffer containing 500 nM REO. The RT-qPCR program was set as follows to generate fluorescence-coded DNA templates: 50°C for 2 min, 95°C for 2 min, followed by 1 cycle of 95°C for 5 s and 60°C for 30 s. After amplification, oligo-dT beads were removed, and the supernatant containing DNA templates and free REO was transferred to 10 μL of streptavidin beads (1 μL of sock beads per reaction) in 2× Binding & Washing Buffer (10 mM Tris-HCl, 1 mM EDTA, 2 M NaCl, 0.1% Tween-20). The solution was incubated on an end-over-end rotator for 5 min to facilitate DNA template capture onto streptavidin beads. Finally, the beads were washed twice with 80 μL of TE buffer (10 mM Tris-HCl, 1 mM EDTA, 0.05% Tween-20) and were ready for downstream ddPCR analysis. For multiplex reactions, the experimental steps remained the same as described above, except that biotinylated LROs and REOs for all targets were used.

Using the Bio-Rad QX200 ddPCR system, streptavidin beads from the previous step were resuspended in 20 μL of ddPCR reaction mixture containing 1.4× ddPCR Supermix, 150 nM forward and reverse universal primers, and 600 nM FAM and HEX LNA probes. Next, 20 μL of the PCR mixture and 70 μL of droplet generation oil were loaded into a DG8 cartridge. The cartridge was sealed with a gasket and placed in the droplet generator to produce approximately 20,000 droplets per well. The generated droplets were then transferred to a 96-well plate, which was sealed with foil film using a plate heat sealer. PCR thermocycling was performed under the following conditions: 95°C for 10 min, followed by 60 cycles of 94°C for 30 s and 60°C for 60 s, then a final step at 98°C for 10 min. After ddPCR, fluorescence intensities of individual droplets were measured in the FAM and HEX channels using the droplet reader.

### Protein detection

For three-protein biomarker detection, 1.5 million antibody-coated beads were used per target per reaction. In experiments conducted with chicken plasma, individual recombinant protein target or mixtures were spiked into 100 μL of 10% chicken plasma and incubated with beads for 15 min to facilitate target capture. Following incubation, beads were washed twice with 80 μL of washing buffer to remove unbound proteins. Beads were then resuspended in PEA buffer containing PEA probes for each target at a final concentration of 5 nM and incubated for 15 min, followed by two additional washes with 80 μL of washing buffer to remove excess PEA probes. Subsequently, beads were resuspended in 10 μL of extension buffer and incubated at 50°C for 5 min, allowing the formation of double-stranded DNA templates on the bead surface. Beads were then washed once prior to the downstream ddPCR reaction as described above. For experiments conducted in human plasma, additional sample preparation was performed due to the high endogenous concentration of CRP. Specifically, plasma samples were diluted 10,000-fold before CRP spike-in or direct testing using the UniQ Buffer. In parallel, plasma samples were diluted 10-fold for IP10 and TRAIL detection using the UniQ Buffer. In the multiplexed assay format, target capture was conducted separately: CRP was processed in one well, while IP10 and TRAIL were processed in a separate well. After the target capture, beads from both wells were combined for subsequent steps. Additionally, when testing human plasma samples, the PEA probe concentration was reduced to 1 nM for CRP and TRAIL to prevent excessive positive droplet formation in ddPCR.

### Oligonucleotide design

The oligonucleotides for mRNA targets were designed as described previously.^28^ Briefly, RT-qPCR primer pairs were designed using IDT PrimerQuest™ Tool, with reverse primers spanning exon-exon junctions (junction primers) to prevent potential false-positive signals from genomic DNA contaminants. The reverse junction primer sequences were adopted for the biotinylated LRO target site, and the sequences immediately upstream were selected for the REO target site. All the RT-qPCR primers, LROs and REOs were designed to have a melting temperature (Tm) of approximately 62°C and a GC content ranging from 35% to 65%. For the JUP, HESX1, and SMARCD3 genes, REO target sites with a GC content higher than 65% were selected due to the limited availability of candidate sequences satisfying our criteria. Additionally, the LROs and REOs were designed to contain universal primer and varying number of TaqMan probe binding sites. For the IFI27 gene, the reverse primer sequence from a previous report^55^ was employed for RT-qPCR and the LRO target site design, followed by the design of the REO target site as described above. For the oligonucleotide sequences used in the PEA probes, each pair included a 5-bp unique hybridization site, in addition to the universal primer and varying number of TaqMan probe binding sites. The 5-bp hybridization sequences were carefully designed and checked using the IDT OligoAnalyzer™ Tool to minimize off-target hybridization and ensure high accuracy of each fluorescence code after PCR amplification.

### Spike-and-recovery test

In the spike-and-recovery test using different sample diluents, 1 ng/mL of each protein target (CRP, IP10, and TRAIL) was spiked into both sample diluent and diluted human plasma (10,000-fold for CRP; 10-fold for IP10 and TRAIL). Non-spiked diluted plasma was included to account for endogenous levels of each target. All samples were processed through the magnetic bead-enhanced PEA workflow, and Ct values were recorded via real-time PCR. The recovery rate was calculated using the following equation:

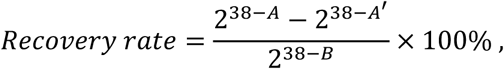

where A, A’, and B correspond to the Ct values obtained from the plasma spike-in sample, the non-spiked plasma sample, and the sample diluent spiked with the target, respectively. Ct values were converted into numbers of starting number of PCR amplicons using 2^38-Ct^, which assumes that 38 is the number of PCR cycles required to bring a single amplicon to the threshold for fluorescence detection.

Following the selection of UniQ Buffer as the optimal sample diluent, its performance was further evaluated using ddPCR, where each target was tested at clinically relevant concentrations, with high, medium, and low spike-in levels as follows: 1 ng/mL, 500 pg/mL, and 250 pg/mL for CRP; 500 pg/mL, 250 pg/mL, and 125 pg/mL for IP10; and 100 pg/mL, 50 pg/mL, and 25 pg/mL for TRAIL. The recovery rate was calculated using the equation:

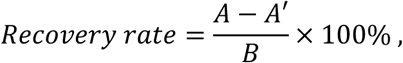

with A, A’, and B corresponding to the ddPCR readouts, denoted in copies/μL, for the plasma spike-in sample, non-spiked plasma sample, and UniQ Buffer spike-in sample, respectively.

### Spiked-and-measured test

The spiked-and-measured test began with standard curve generation using 2-fold serial dilutions of each target protein spiked into UniQ Buffer: CRP (1000– 31.25 pg/mL), IP10 (500–15.625 pg/mL), and TRAIL (100–3.125 pg/mL). Standard curves were plotted by normalizing ddPCR readouts at each concentration to the signal obtained from the no-target control and plotting these values against the input target concentrations. Next, 2-fold serial dilutions of each target protein were spiked into diluted human plasma: CRP into 10,000-fold diluted plasma (800–50 pg/mL), and IP10 and TRAIL into 10-fold diluted plasma (IP10: 400– 25 pg/mL; TRAIL: 80–5 pg/mL). The ddPCR outputs were used to calculate the measured target concentrations, which were then plotted against the spiked concentrations with linear regression fitting, with a slope near 1 and minimal intercept indicating high quantification accuracy. In the multiplexed assay format, the spiked-and-measured test was performed using target mixtures at varying concentrations as above, along with antibody-coated beads and PEA probes in the same well. For plasma spike-in conditions, the target capture step for CRP detection was conducted separately from IP10 and TRAIL, while all subsequent steps were performed together.

### Codetection of mRNA and protein biomarkers in a single ddPCR reaction

Beads carrying fluorescence-coded DNA templates from mRNA and protein targets were pooled into a single ddPCR reaction containing 1.4× ddPCR Supermix, 150 nM forward and reverse universal primers, and 600 nM FAM and HEX LNA probes. The reaction mixture was then processed through droplet generation, ddPCR amplification, and droplet reading. While mRNA quantification was performed directly from ddPCR readouts, protein quantification required the generation of standard curves. Therefore, when detecting proteins from plasma samples, a standard curve was constructed for each target in parallel within a multiplexed assay format using UniQ Buffer for every experimental run.

### Low-volume blood sample processing

Human whole blood was processed through both RNA extraction and plasma separation to enable mRNA and protein measurement. For RNA stabilization, blood was mixed with Tempus solution at a 1:2 volume ratio in a 1.5 mL tube to stabilize RNA. The mixture was vortexed vigorously for 15 seconds and stored at 4°C until RNA extraction. RNA was extracted using the Tempus™ Spin RNA Isolation Kit according to the manufacturer’s protocol (no DNase treatment) with two modifications: the volume of 1× PBS used was adjusted to match the volume of blood added, and the final elution volume was reduced to 50 μL instead of 100 μL. After extraction, RNA yield was measured using the Qubit RNA HS Assay kit. For plasma separation, fresh blood was centrifuged at 2000 xg for 15 min. Plasma quality was assessed by measuring the absorbance at 540 nm using NanoDrop.

### Data analysis

Each multiplexed assay result, whether measuring mRNA, protein, or both, was analyzed using custom-developed Python scripts incorporating packages such as NumPy, pandas, matplotlib, SciPy, and the GMM from the scikit-learn machine learning library. Briefly, fluorescence intensity data at both FAM and HEX channels for individual droplets from each ddPCR reaction were imported from CSV files. GMM was applied to identify negative droplets, outliers, and target-specific clusters (**Figure S15**). To ensure robust clustering for each mRNA target, especially low-abundance targets such as HESX1, EBI3, and JUP, ddPCR signals from pooled synthetic DNA templates were used to enable reliable identification of each clusters. Employing Poisson distribution statistics in ddPCR, the target concentration (C) in each cluster was calculated as follows:

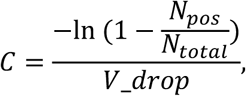

where *N_pos_* is the number of positive droplets, *N_Total_* is total number of droplets read by the droplet reader, and *V_drop* is the volume of individual droplets (1 nL) generated on Bio-Rad QX200 ddPCR system.

### RT-qPCR

The single-plex RT-qPCR reaction was performed in a 10-μL reaction mixture containing 1× Lyoready RT-qPCR master mix, 100 nM target-specific primers, 1× Evagreen dye, and 1 μL human total RNA. The reaction mix was incubated in a Bio-Rad CFX96 Touch Real-Time PCR Detection System (Bio-Rad, Hercules, CA) at 50 °C for 10 min, 95 °C for 2 min, followed by 50 cycles of 95 °C for 5 s, 60 °C for 20 s with the fluorescence measured at 60 °C in each cycle.

### ELISA

ELISAs were performed using commercial kits according to the manufacturers’ instructions. Briefly, 20 μL and 50 μL of human plasma (the maximum input volume) were added to capture antibody-coated wells for the IP10 and TRAIL assays, respectively. For CRP measurement, plasma samples were diluted 10,000-fold prior to testing. Standard curves were generated by testing two-fold serial dilutions of the standard proteins in the respective sample diluent buffer. Following sample incubation and washing steps, a peroxidase-conjugated detection antibody was added to form immune complexes with the target proteins. After extensive washing, substrate solution was added to each well to allow color development. Optical density was measured at 450 nm using a plate reader, with wavelength correction to minimize the effects of optical imperfections. Target protein concentrations in plasma samples were calculated based on the corresponding standard curves.

## Data Availability

All data produced in the present study are available upon reasonable request to the authors.

## Acknowledgments

The authors are grateful for the financial support from the National Institutes of Health (R01CA260628, R33CA272321, R01AI183336, and R01AI181217).

