## Supplementary information for "A Unified Multiplex Digital PCR-based Platform for Multi-class Biomarker Quantification from Low-volume Blood"

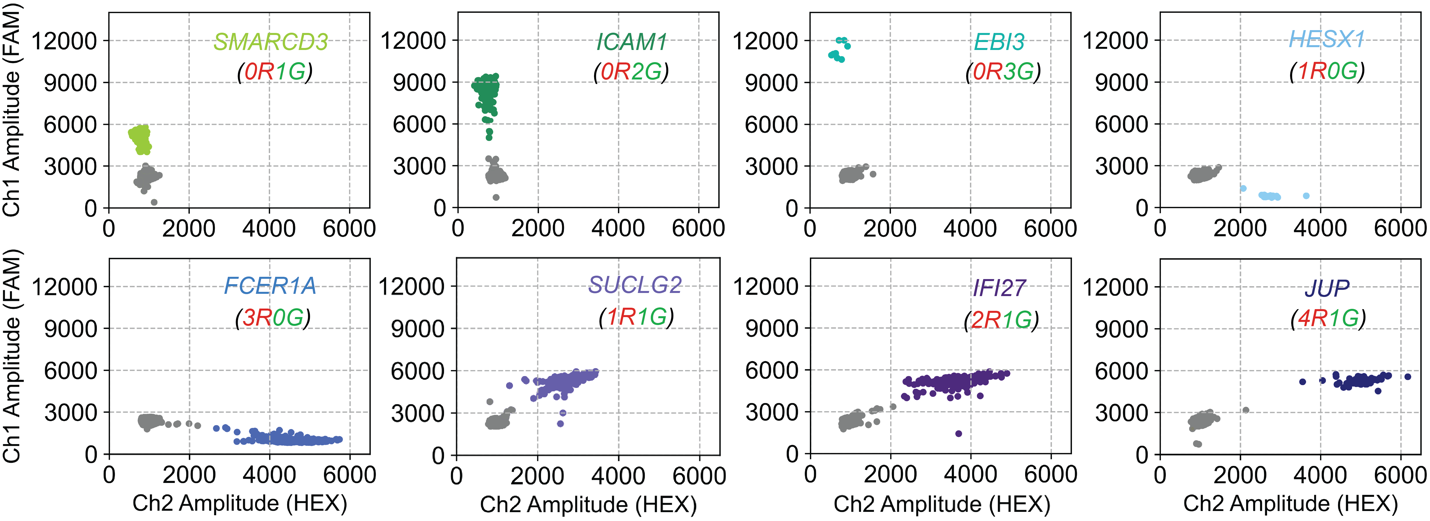

**Figure S1**. Two-dimensional amplitude plots of singleplex mRNA assays following fluorescence-coding ddPCR. Each datapoint represents a single droplet. Gray droplets indicate negatives, while colored droplets correspond to specific fluorescence-coded DNA targets.

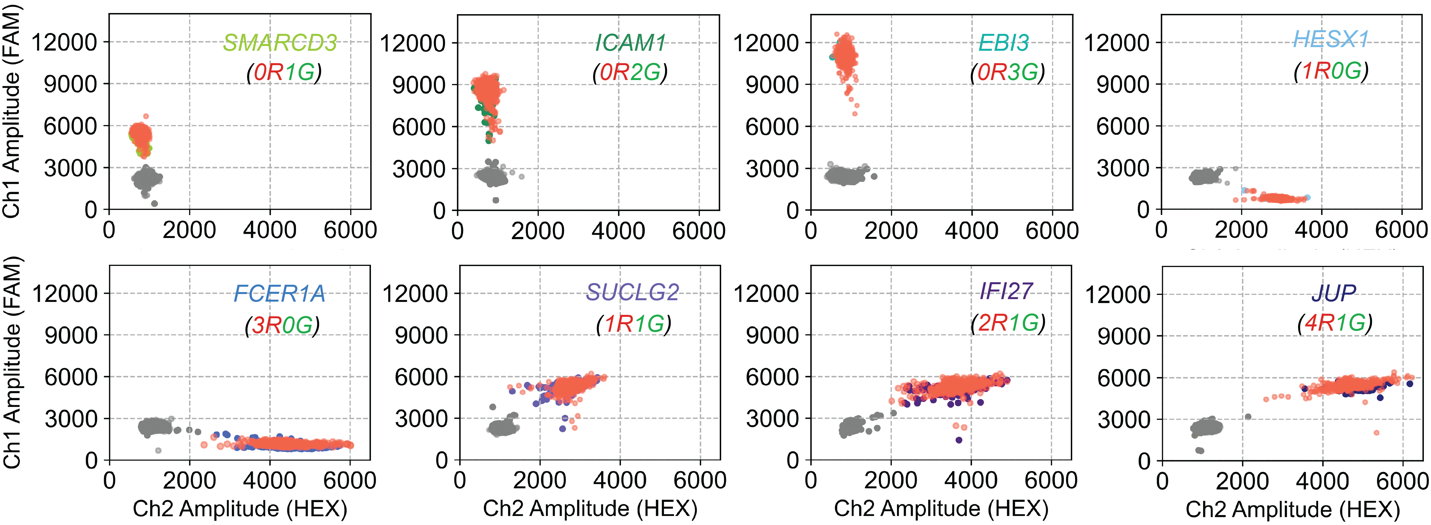

**Figure S2.** Synthetic fluorescence-coded DNA templates, designed to match the sequences of mRNA-derived amplicons for each target, produced identical fluorescence signatures following ddPCR (in red).

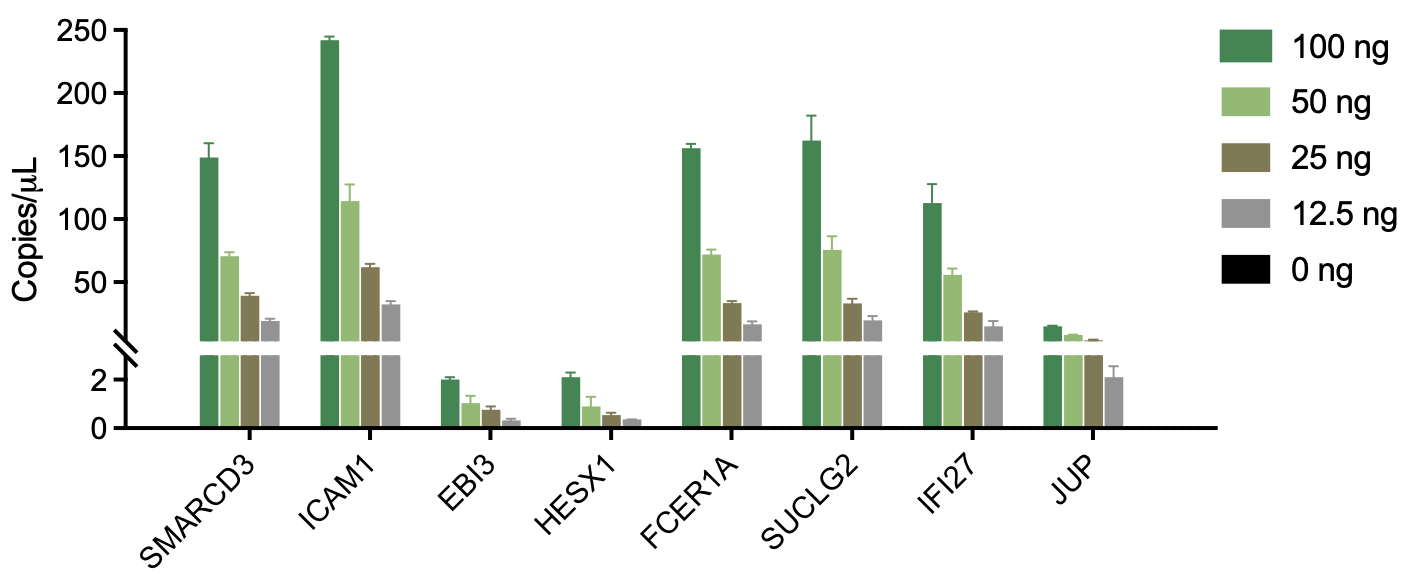
**Figure S3.** Bar plots showing ddPCR signal outputs from singleplex mRNA assays using varying amounts of human total RNA input. Each assay produced distinct, input-dependent signals with clean background. Bars represent mean ± standard deviation from three replicates.

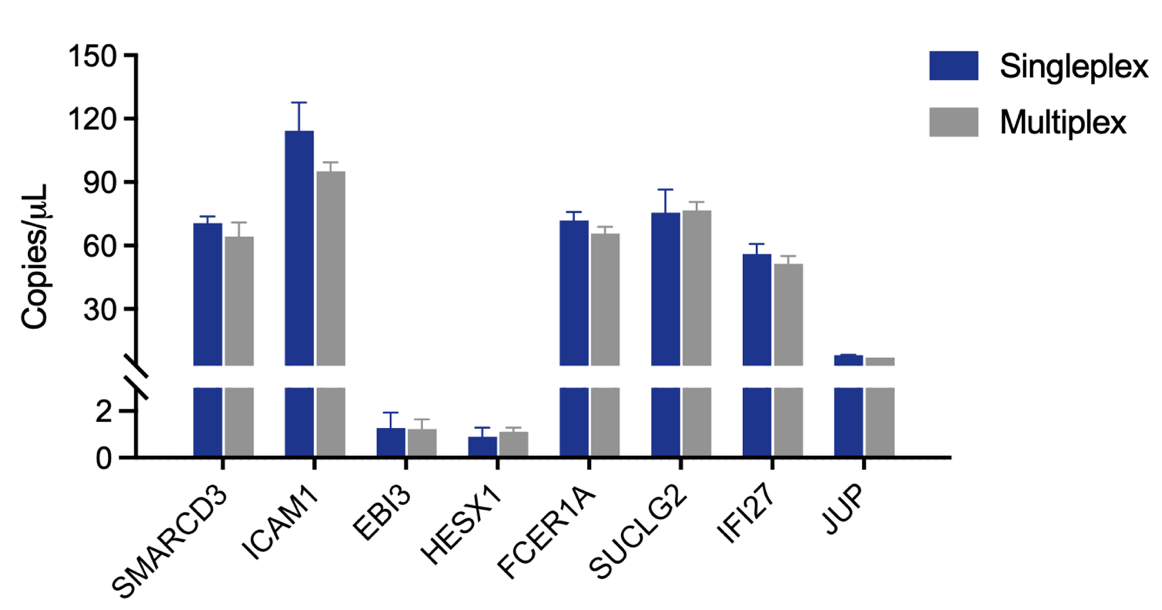

**Figure S4.** Comparison of ddPCR readouts for each mRNA target obtained from the multiplex assay versus the corresponding singleplex assays. Reactions were performed with 50 ng of human total RNA input. Data represent mean ± standard deviation from three replicates.

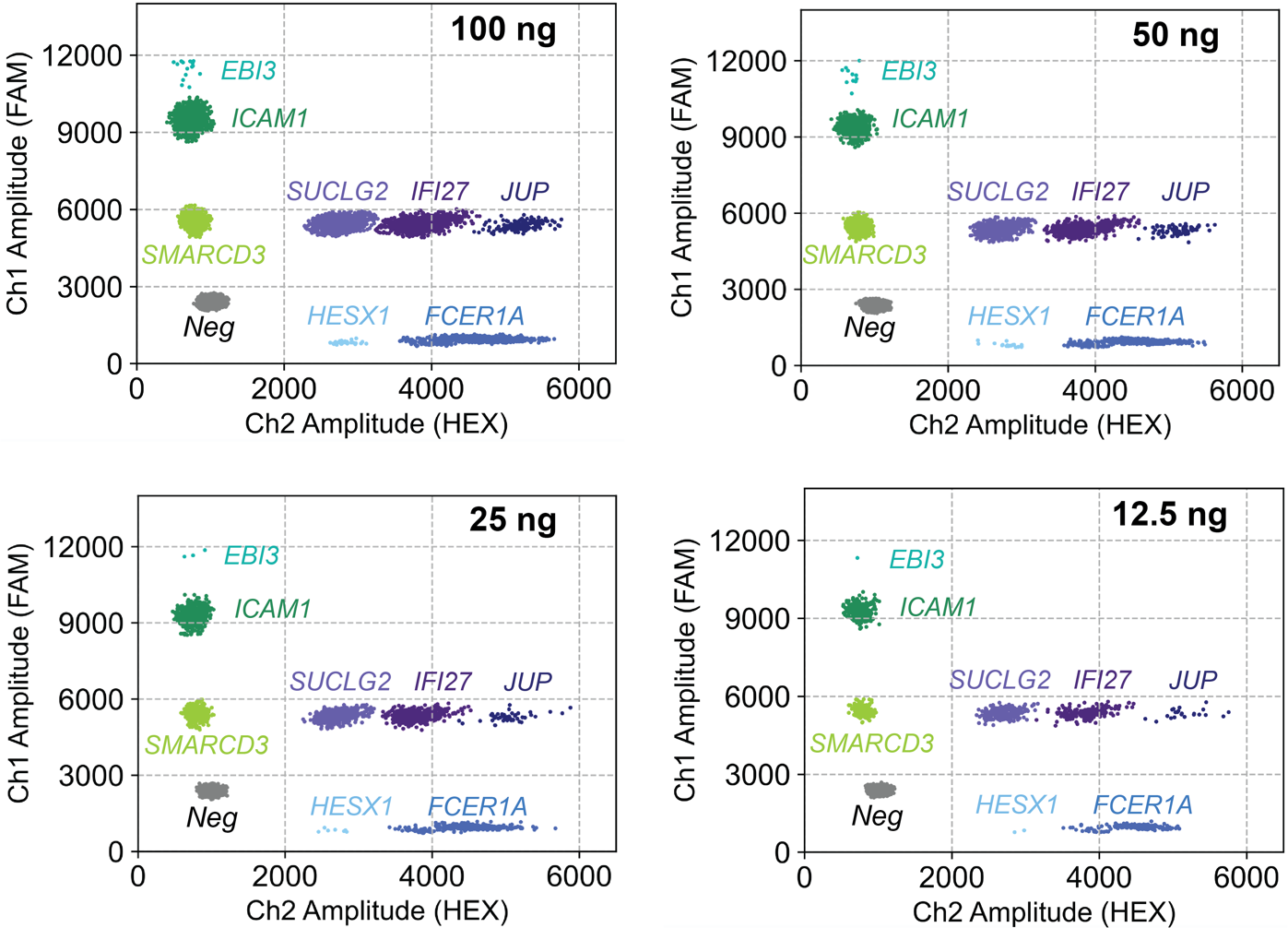

**Figure S5.** Two-dimensional FAM-versus-HEX amplitude plots from multiplex ddPCR reactions using varying amounts of human total RNA. All reactions consistently show eight target-specific droplet populations along with a distinct negative population.

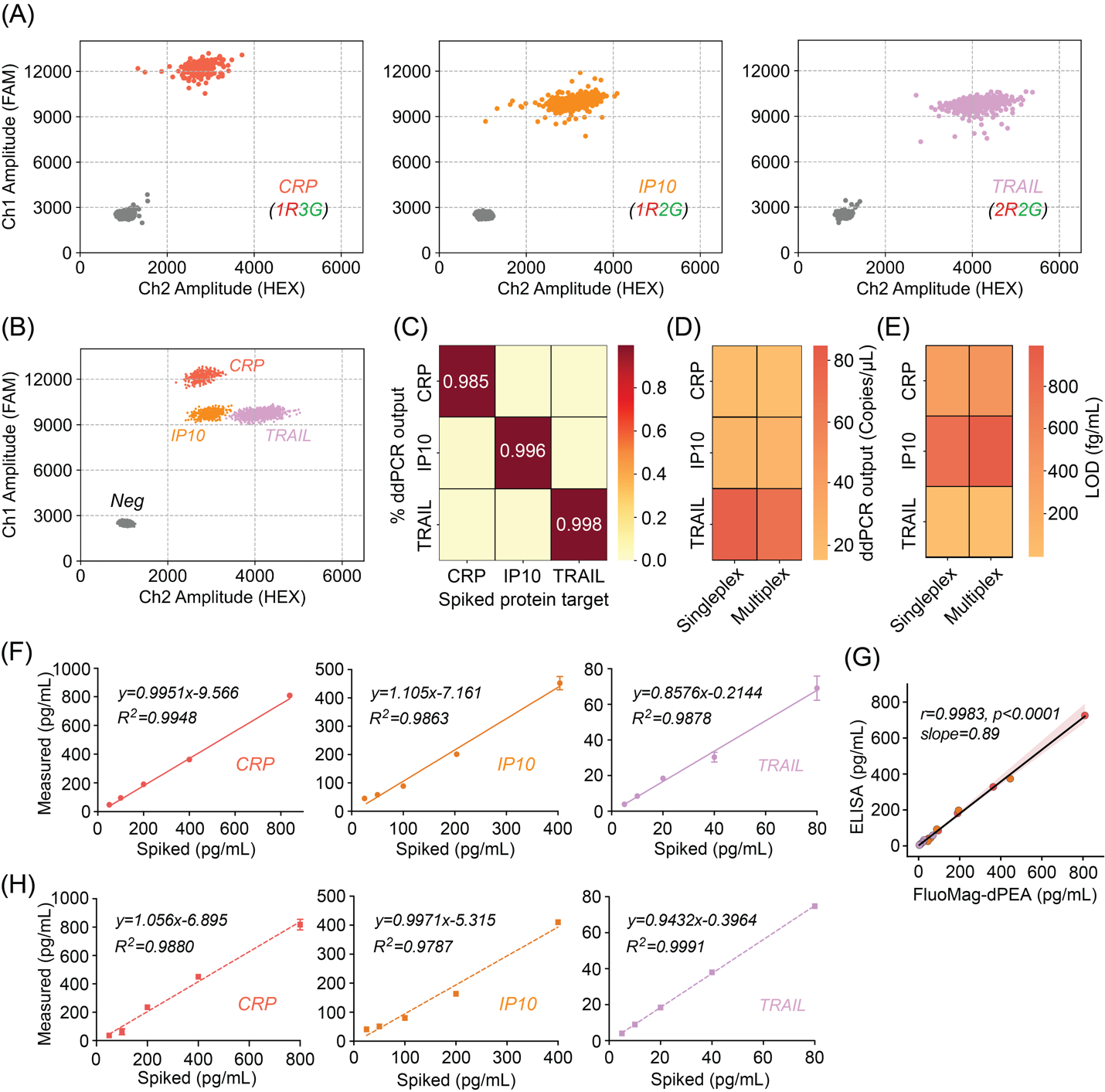

**Figure S6.** Two-dimensional amplitude plots of singleplex protein assays following fluorescence-coded ddPCR. Each datapoint represents an individual droplet. Gray droplets indicate negatives, while colored droplets correspond to the converted DNA targets with distinct fluorescence-codes.

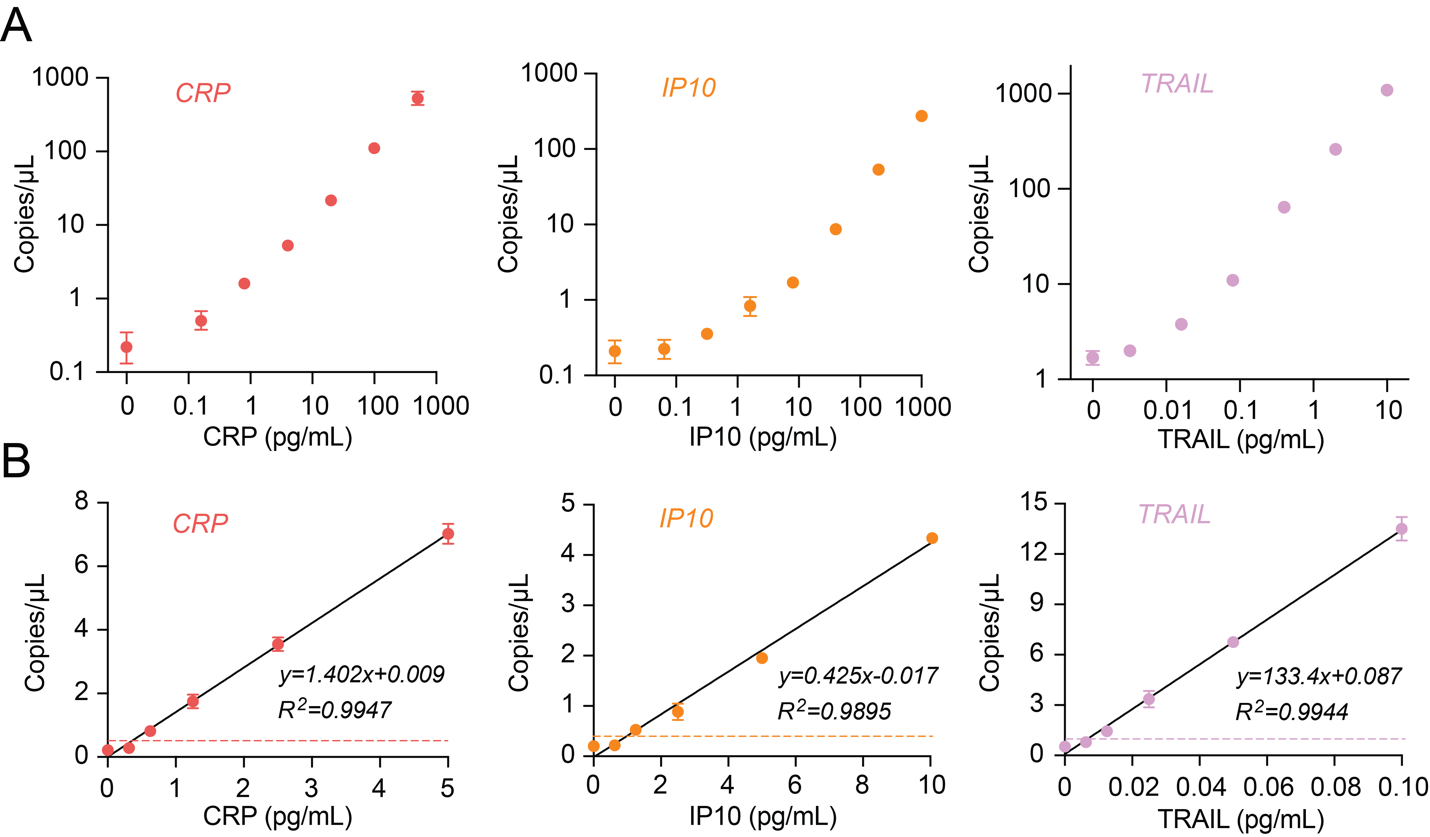

**Figure S7.** Singleplex validation of each protein target in chicken plasma. (A) Serially diluted recombinant proteins spiked in 10% chicken plasma were tested, and ddPCR outputs were plotted against target concentrations on a log–log scale to assess dynamic range. (B) Assay response of each target in the sub-nanogram or femtogram ranges demonstrate linearity. LODs were calculated by extrapolating the concentration corresponding to the background signal plus three times its standard deviation. Dashed lines indicate the ddPCR signals at the LOD for each target. Error bars represent data from two replicates.

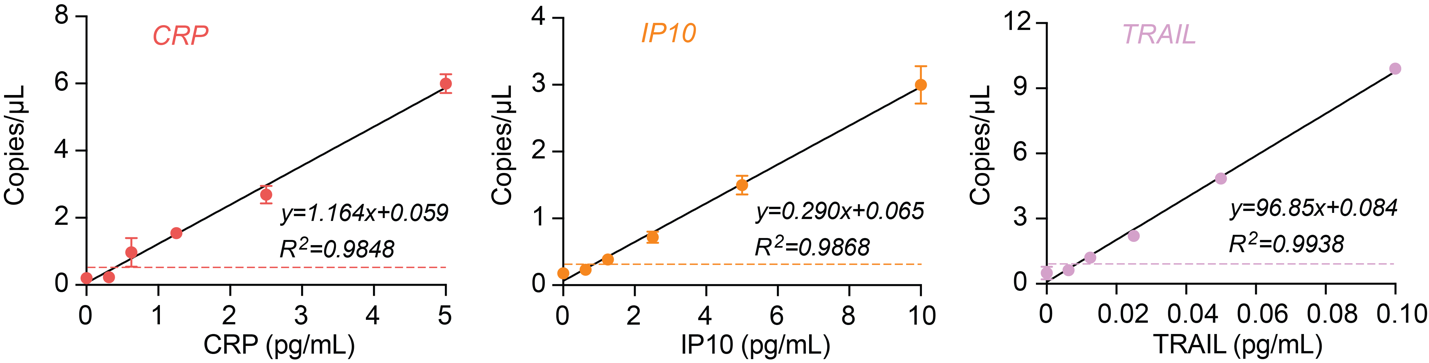

**Figure S8.** Assay response curves for each target in a multiplex reaction. Dashed lines indicate the ddPCR signal corresponding to the LOD for each target. Error bars represent data from two replicates.

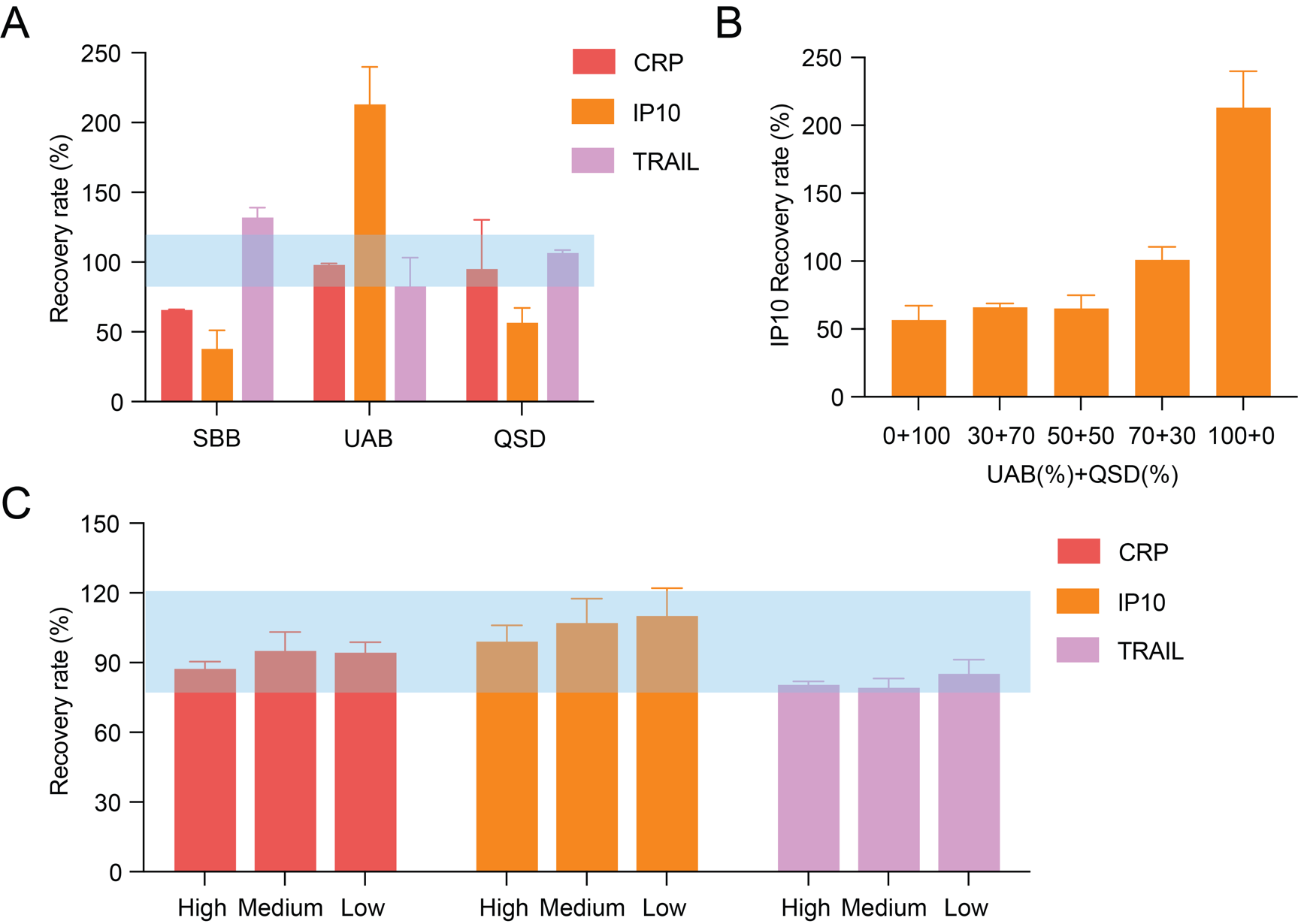

**Figure S9.** Spike-and-recovery analysis (via real-time qPCR) for each protein target in three commercially available sample diluents. The blue shaded area indicates the 80%–120% recovery range, representing acceptable accuracy for quantification. Data are presented as mean ± standard deviation from three replicates.

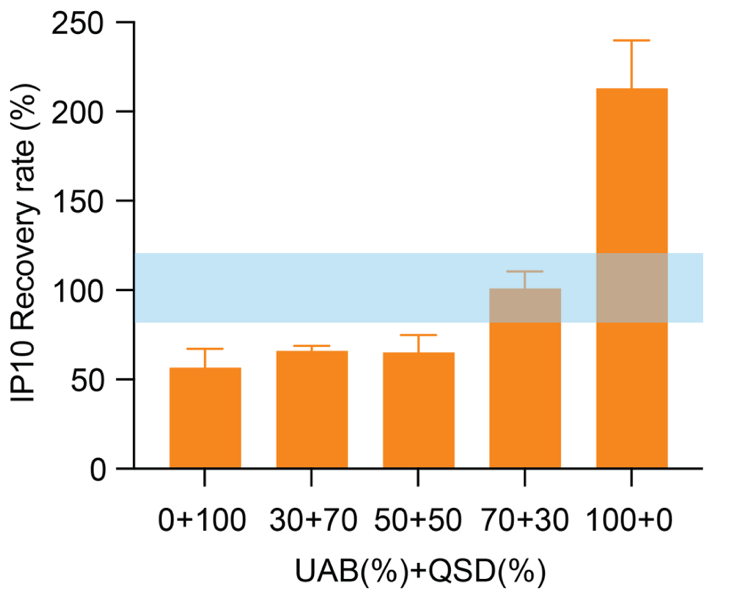

**Figure S10.** Spike-and-recovery tests (via real-time qPCR) of IP-10 across different sample diluent compositions (n ≥ 2) containing varying proportions of UAB and QSD.

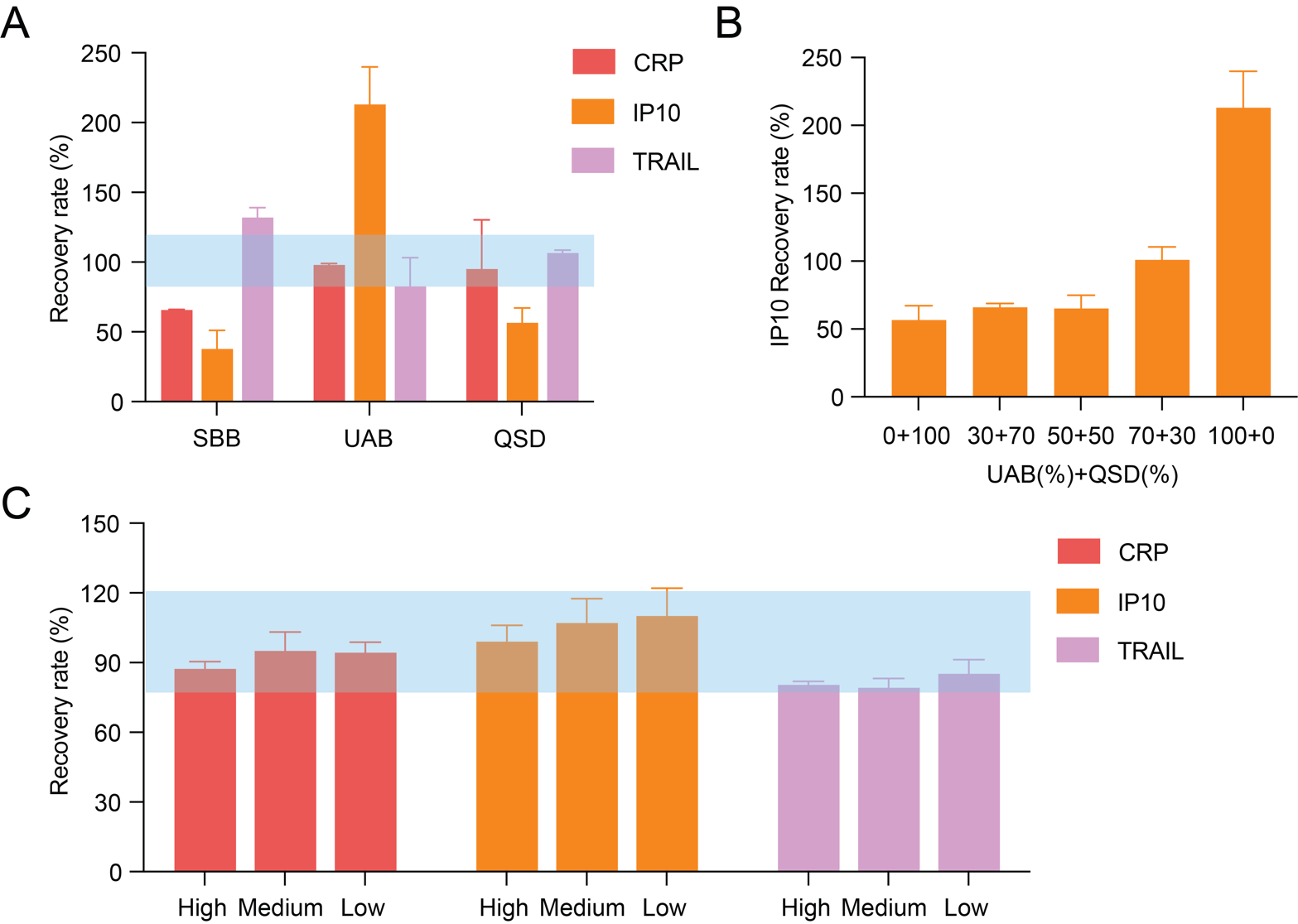

**Figure S11.** Spike-and-recovery analysis via ddPCR for each protein target in UniQ Buffer with varying spiked input concentrations. The blue shaded area indicates the recovery range of 80%–120%. Data are presented as mean ± standard deviation from three replicates.

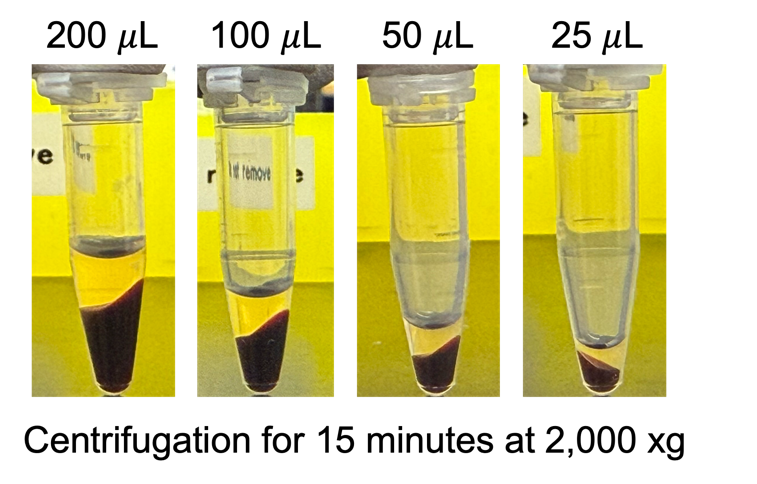

**Figure S12.** Images of plasma separation from human whole blood following centrifugation, showing a clear upper plasma layer and a darker lower layer of cellular components. Effective separation was achieved across all tested blood volumes, from 200 μL down to 25 μL.

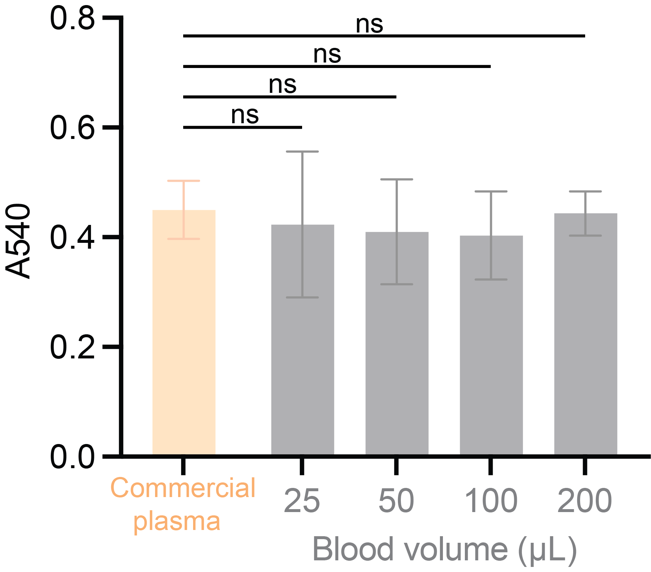

**Figure S13.** Spectrophotometric comparison of absorbance at 540 nm between separated plasma and commercially sourced plasma. No significant difference was observed (ns, Student’s t-test), indicating high purity of plasma obtained from low-volume blood samples. Bars represent the mean ± standard deviation from three replicates.

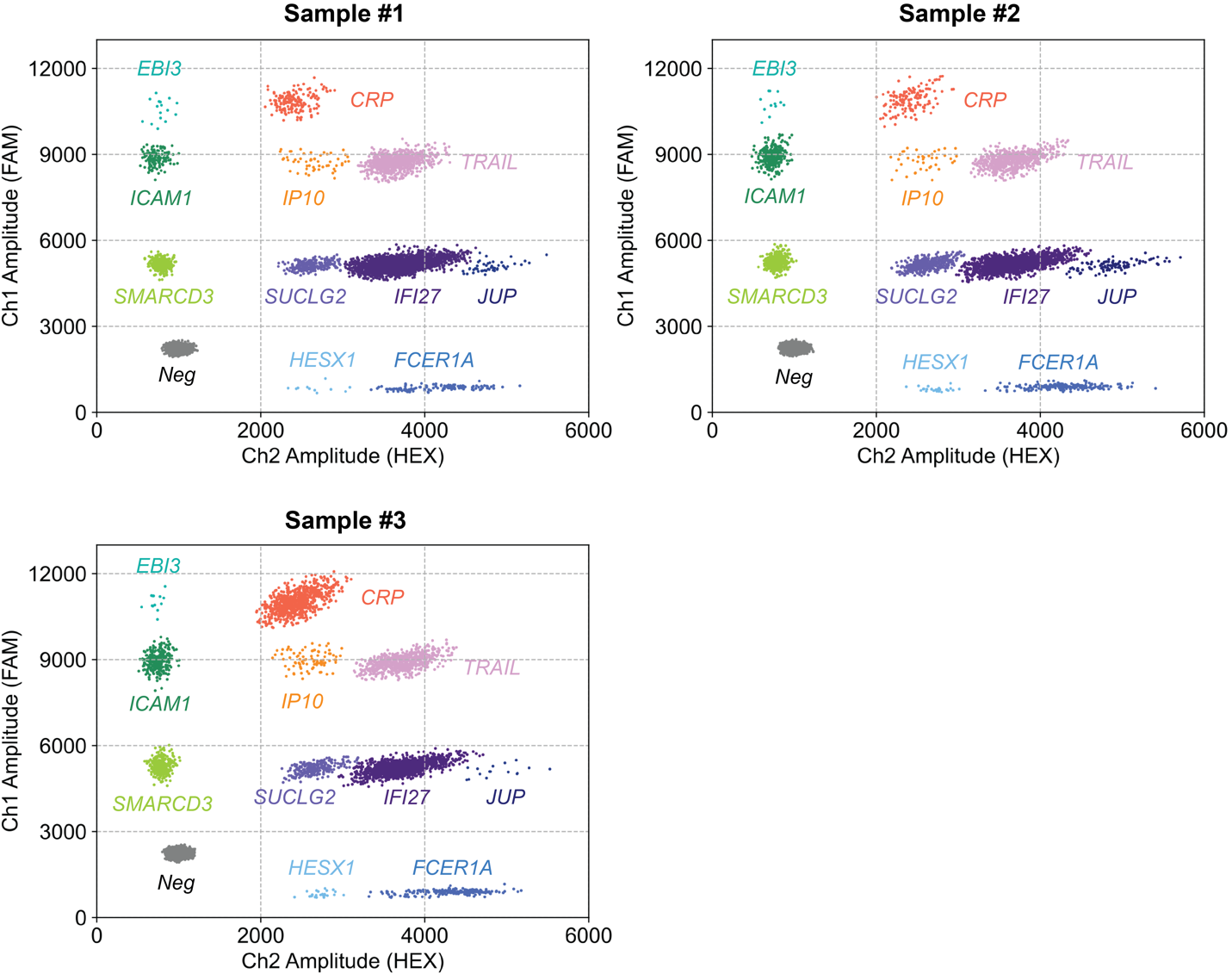

**Figure S14.** Two-dimensional amplitude plots from one-pot ddPCR reactions using 10 μL of blood for mRNA detection and 25 μL for protein detection across three independent blood batches.

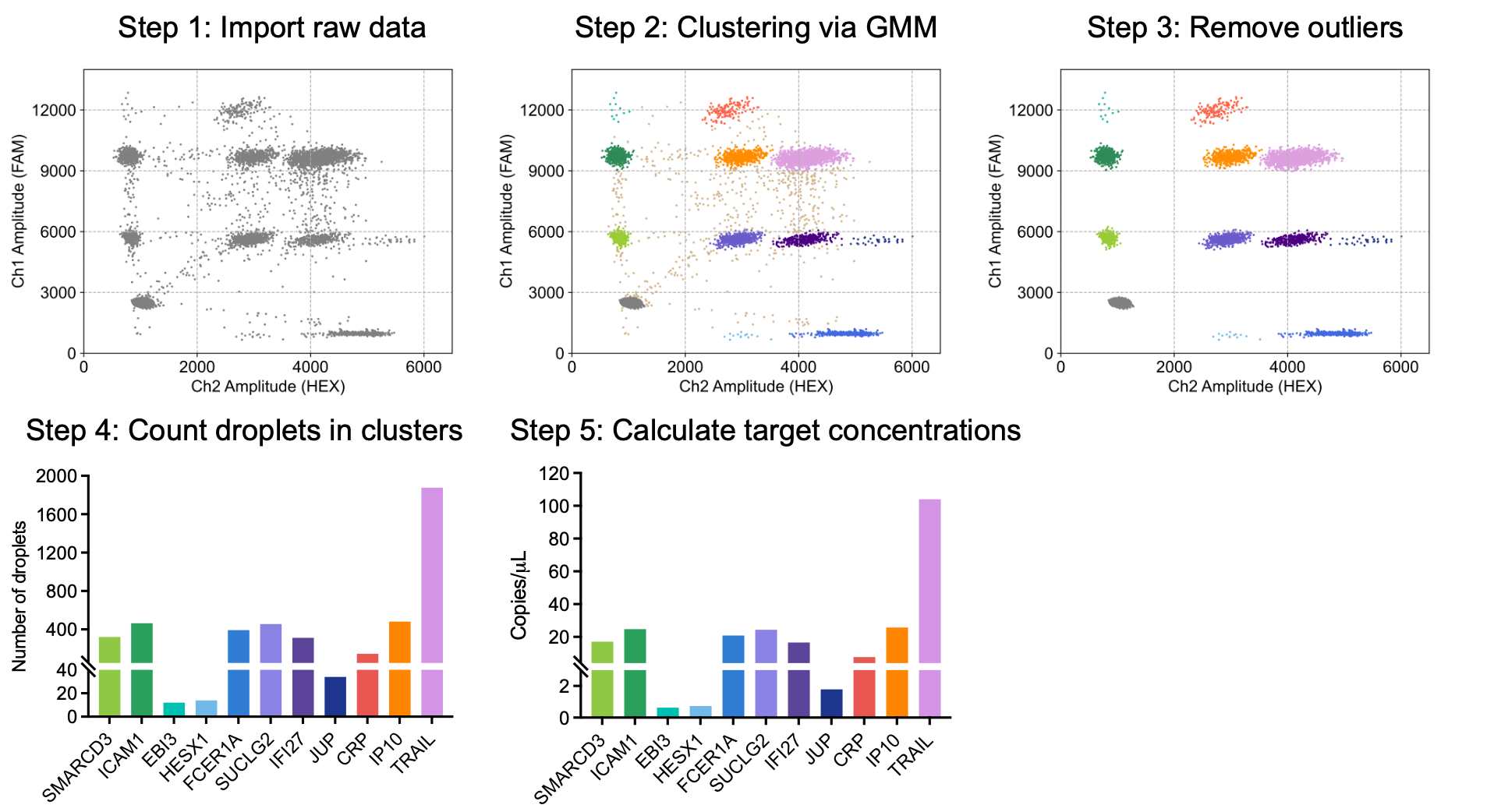

**Figure S15.** Data analysis workflow implemented in Python. GMM was applied to the raw ddPCR data to identify target-specific clusters, negative droplets, and outliers. Within each target-specific cluster, positive droplets were counted, and ddPCR signal outputs were calculated using Poisson distribution statistics.

**Table S1**. Sequences of LROs, REOs, PEA probe oligonucleotides, universal primers, and TaqMan probes.

| **Item** | **Sequence (5’ to 3’)** |
| --- | --- |
| SMARCD3 LRO | /5BiosG/TATATATACAACACCCCAACATCTTCGAGCTGGATCTTGCAGTAGAGTAG |
| SMARCD3 REO | TGGAAGTTCACCGGCGTCATCGACGAACGGTGCAGGAGGCCGTCAGTCG |
| ICAM1 LRO | /5BiosG/TATATATACAACACCCCAACATCTTCGATGGAGTCCAGTACACGGT |
| ICAM1 REO | TGGAAGTTCACCGGCGTCATCGACGAACGGTGCTTTCGACGAACGGTGCAGTCAACAGCTAAAACCTTCCTC |
| EBI3 LRO | /5BiosG/TATATATACAACACCCCAACATCTTCGAGGTCGGGCTTGATGATGT |
| EBI3 REO | TGGAAGTTCACCGGCGTCATCGACGAACGGTGCTTTCGACGAACGGTGCTTTCGACGAACGGTGCTCGTGCCTTTCATAACAGAGC |
| HESX1 LRO | /5BiosG/TATATATACAACACCCCAACATCTTCGACGCTCGACGTAGCAAGTTACCATCTTTCCCTGATGA |
| HESX1 REO | TGGAAGTTCACCGGCGTCATGGCAGACACCTGCAGC |
| FCER1A LRO | /5BiosG/TATATATACAACACCCCAACATCTTCGACGCTCGACGTAGCTTTCGCTCGACGTAGCTTTCGCTCGACGTAGCCTTCTCACGCGGAGCTTT |
| FCER1A REO | TGGAAGTTCACCGGCGTCATGAGCCCCTCAACATTACTGTAATA |
| SUCLG2 LRO | /5BiosG/TATATATACAACACCCCAACATCTTCGACGCTCGACGTAGCATGGCTTCAACCTTAGGATCAG |
| SUCLG2 REO | TGGAAGTTCACCGGCGTCATCGACGAACGGTGCCAAGCATTCAAATTGCTCACAG |
| IFI27 LRO | /5BiosG/TATATATACAACACCCCAACATCTTCGACGCTCGACGTAGCTTTCGCTCGACGTAGCACATCATCTTGGCTGCTATGG |
| IFI27 REO | TGGAAGTTCACCGGCGTCATCGACGAACGGTGCGGAATCGCCTCGTCCT |
| JUP bLRO | /5BiosG/TATATATACAACACCCCAACATCTTCGACGCTCGACGTAGCTTTCGCTCGACGTAGCTTTCGCTCGACGTAGCTTTCGCTCGACGTAGCTCCTCACACCATCCGTGTA |
| JUP REO | TGGAAGTTCACCGGCGTCATCGACGAACGGTGCGGCACACAGCAGCCC |
| CRP probe A | /5ThioMC6D/TTTTTTTTACAACACCCCAACATCTTCGACGCTCGACGTAGCGGATC |
| CRP probe B | /5ThioMC6D/TTTTTTTGGAAGTTCACCGGCGTCATCGACGAACGGTGCTTTCGACGAACGGTGCTTTCGACGAACGGTGCGATCC |
| IP10 probe A | /5ThioMC6D/TTTTTTTTACAACACCCCAACATCTTCGACGCTCGACGTAGCGCAGG |
| IP10 probe B | /5ThioMC6D/TTTTTTTGGAAGTTCACCGGCGTCATCGACGAACGGTGCTTTCGACGAACGGTGCCCTGC |
| TRAIL probe A | /5ThioMC6D/TTTTTTTTACAACACCCCAACATCTTCGACGCTCGACGTAGCTTTCGCTCGACGTAGCTGCCA |
| TRAIL probe B | /5ThioMC6D/TTTTTTTGGAAGTTCACCGGCGTCATCGACGAACGGTGCTTTCGACGAACGGTGCTGGCA |
| Forward primer | TACAACACCCCAACATCTTCGA |
| Reverse primer | GGAAGTTCACCGGCGTCAT |
| FAM probe | /5FAM/CGaCGaAcGgtGC/3BHQ1/* |
| HEX probe | /5HEX/CGcTcGAcGtAgC/3IABkFQ/* |

The underline bases represent hybridization sites

* “a”, “c”, “g”, “t” indicates locked nucleic acid

**Table S2**. Synthetic fluorescence-coded DNA template sequences corresponding to each mRNA target.

| **Item** | **Sequence (5’ to 3’)** |
| --- | --- |
| SMARCD3 | TGGAAGTTCACCGGCGTCATCGACGAACGGTGCAGGAGGCCGTCAGTCG**CTACTTCTACTGCAAGATCCAGCTCGAAGATGTTGGGGTGTTGTAA** |
| ICAM1 | TGGAAGTTCACCGGCGTCATCGACGAACGGTGCTTTCGACGAACGGTGCAGTCAACAGCTAAAACCTTCCTC**ACCGTGTACTGGACTCCATCGAAGATGTTGGGGTGTTGTAA** |
| EBI3 | TGGAAGTTCACCGGCGTCATCGACGAACGGTGCTTTCGACGAACGGTGCTTTCGACGAACGGTGCTCGTGCCTTTCATAACAGAGC**ACATCATCAAGCCCGACCTCGAAGATGTTGGGGTGTTGTAA** |
| HESX1 | TGGAAGTTCACCGGCGTCATGGCAGACACCTGCAGC**TCATCAGGGAAAGATGGTAACTTGCTACGTCGAGCGTCGAAGATGTTGGGGTGTTGTAA** |
| FCER1A | TGGAAGTTCACCGGCGTCATGAGCCCCTCAACATTACTGTAATA**AAAGCTCCGCGTGAGAAGGCTACGTCGAGCGAAAGCTACGTCGAGCGAAAGCTACGTCGAGCGTCGAAGATGTTGGGGTGTTGTAA** |
| SUCLG2 | TGGAAGTTCACCGGCGTCATCGACGAACGGTGCCAAGCATTCAAATTGCTCACAG**CTGATCCTAAGGTTGAAGCCATGCTACGTCGAGCGTCGAAGATGTTGGGGTGTTGTAA** |
| IFI27 | TGGAAGTTCACCGGCGTCATCGACGAACGGTGCGGAATCGCCTCGTCCT**CCATAGCAGCCAAGATGATGTGCTACGTCGAGCGAAAGCTACGTCGAGCGTCGAAGATGTTGGGGTGTTGTAA** |
| JUP | TGGAAGTTCACCGGCGTCATCGACGAACGGTGCGGCACACAGCAGCCC**TACACGGATGGTGTGAGGAGCTACGTCGAGCGAAAGCTACGTCGAGCGAAAGCTACGTCGAGCGAAAGCTACGTCGAGCGTCGAAGATGTTGGGGTGTTGTAA** |

(1) Underline bases represent hybridization sites to mRNA targets

(2) Non-bolded bases are identical to the corresponding REO sequences for each mRNA target

(3) Bolded bases are reverse complementary to the corresponding LRO sequences for each mRNA target

**Table S3**. ANCOVA F-test showing the difference in slope and intercept between singleplex and multiplex linear fitting curves for each mRNA target.

| **Target** | **Slope (singleplex/multiplex)** | **Intercept (singleplex/multiplex)** | **Slope comparison by P value*** | **Intercept comparison by P value*** | **Pooled slope^⊥^** | **Pooled intercept^⊥^** |
| --- | --- | --- | --- | --- | --- | --- |
| SMARCD3 | 1.449/1.449 | 1.080/-0.765 | 0.999 | 0.301 | 1.449 | 0.157 |
| ICAM1 | 2.347/2.318 | 0.026/-4.848 | 0.762 | 0.059 | 2.332 | -2.411 |
| EBI3 | 0.018/0.019 | 0.176/-0.019 | 0.570 | 0.053 | 0.019 | 0.079 |
| HESX1 | 0.023/0.023 | -0.014/0.032 | 0.813 | 0.438 | 0.023 | 0.009 |
| FCER1A | 1.609/1.545 | -5.903/-3.368 | 0.186 | 0.770 | 1.577 | -4.636 |
| SUCLG2 | 1.660/1.629 | -5.128/0.319 | 0.800 | 0.318 | 1.645 | -2.404 |
| IFI27 | 1.131/1.170 | -0.561/2.158 | 0.703 | 0.187 | 1.151 | 0.799 |
| JUP | 0.143/0.146 | 0.380/-0.232 | 0.783 | 0.062 | 0.144 | 0.074 |

* P > 0.05 means the singleplex and multiplex linear curves have indistinguishable slopes or intercepts for that target.

^⊥^ The single slope or intercept that can best describe both singleplex and multiplex datasets.

**Table S4**. RT-qPCR primer sequences for each mRNA target.

| **Item** | **Sequence (5’ to 3’)** |
| --- | --- |
| SMARCD3-FWD | CCTCAAGGTGATGACAGATGTAG |
| SMARCD3-REV | GCTGGATCTTGCAGTAGAAGTAG |
| ICAM1-FWD | GAACTGAGCAATGTGCAAGAAG |
| ICAM1-REV | TGGAGTCCAGTACACGGT |
| EBI3-FWD | GATGTCCAGCTGTTCTCCAT |
| EBI3-REV | GGTCGGGCTTGATGATGT |
| HESX1-FWD | GAATCTTAGGACTGGACCAGAAG |
| HESX1-REV | AAGTTACCATCTTTCCCTGATGA |
| FCER1A-FWD | CAGTTGAAGACAGTGGAACCT |
| FCER1A-REV | CTTCTCACGCGGAGCTTT |
| SUCLG2-FWD | GGATCTTGGAGGTGGTGTAAAG |
| SUCLG2-REV | ATGGCTTCAACCTTAGGATCAG |
| IFI27-FWD | GCTACAGTTGTGATTGGAGGAG |
| IFI27-REV | ACATCATCTTGGCTGCTATGG |
| JUP-FWD | GCCTCGTCCAACTGCTG |
| JUP-REV | TCCTCACACCATCCGTGTA |
